# An fMRI Phenotype of Reward in Effort Expenditure in the VTA and Dorsal Raphe: A Longitudinal Study of Depression under Escitalopram Treatment

**DOI:** 10.64898/2026.07.24.26358864

**Authors:** Mirna Hajrić, Elisa Sittenberger, Lisa Dommes, Julia E. Bosch, Petra Beschoner, Franziska Geiser, Julia C. Stingl, Roberto Viviani

## Abstract

Anhedonia and motivational impairment are central features of major depressive disorder, yet the neural processes linking reward information processing to sustained goal-directed behaviour remain insufficiently understood. Using functional magnetic resonance imaging, we examined representations of reward levels during sustained effort expenditure (a construct in the RDoC framework) as a distinct component of reward processing in depressive patients under escitalopram treatment. Twenty-eight unmedicated patients with a current depressive episode and forty-three healthy control participants underwent fMRI scanning at two time points three weeks apart; patients started escitalopram immediately after the first measurement. Across participants, sustained effort expenditure engaged midbrain reward circuitry (ventral tegmental area, VTA), even in the absence of immediate reinforcement. Here, we observed a reward level x session x patient group interaction, driven by larger activity in patients after treatment. A targeted analysis also revealed changes in activity of the dorsal raphe nucleus (DRN) in patients. By isolating neural mechanisms supporting sustained goal-directed behaviour, the study identifies novel and clinically relevant imaging phenotypes of reward processing and their change after three weeks of treatment, representing a potential pathway of escitalopram treatment in depression.

## 1 Introduction

Amotivation and anhedonia represent core disabling features of major depressive disorder (MDD), a centrality recognized since the earliest diagnostic criteria ^1^. Because of the considerable evidence in animal and human studies on the key role of dopamine in motivation and reward learning, an important strand of research has focused on altered dopaminergic function in depression (for a review, see Pizzagalli, 2022 ^2^). However, selective serotonin reuptake inhibitors (SSRIs), the mainstay of the pharmacology of depression, act by enhancing the activity of the serotonergic system, which has been traditionally viewed as being functionally opposed to the dopaminergic system ^3–5^.

Several possibilities exist to explain this conundrum. One is that the evidence for the opponency between these two systems has been mixed ^6^, a possible consequence of the heterogeneous nature of dorsal raphe organization ^7,8^. Another is to note that while symptoms such as anhedonia are among those least improved by SSRIs ^9,10^, anhedonia is viewed in psychopathological models as not limited to diminished pleasure but also involving disruptions across several stages of reward processing ^2,11^. Hence, SSRIs may differentially affect the components of the motivational/hedonic symptom complex, improving some facets but not others. This underscores the need for a comprehensive mapping of the processes that contribute to the motivational/hedonic components of depression to assess the possible impact of depression treatment. In parallel, the possible role of the serotonergic system in the components of the motivational/hedonic symptom complex needs to be reassessed. Recent animal studies show the existence of a feedback loop between raphe nuclei and the reward processing network where the serotonergic system is thought to have a driving role (for a review, see Koolschijn et al., 2024 ^12^).

Reward processing involves three main phases: (a) a motivational phase (i.e., the incentive and motivation to reach a goal), (b) an intermediate phase (i.e., the recruitment of effort during goal pursuit), and (c) a valuation phase (i.e., experiencing pleasure when consuming the reward ^11^). While the motivational and valuation phases have been extensively researched, less is known about the intermediate phase. In the Research Domain Criteria (RDoC), the computation of the value of a reward while considering both its magnitude and the perceived costs of the cognitive effort required to obtain it is characterized as “effort expenditure”, a subconstruct within the “reward valuation” construct in the positive valence system^13^. This construct was based on the Effort Expenditure for Rewards Task (EEfRT), developed from paradigms in laboratory animals showing the invigorating effects of dopamine ^14^. Its relevance is supported by evidence from behavioural studies. In the EEfRT, individuals with higher anhedonia and patients with MDD show reduced willingness to exert effort for potential rewards and reduced sensitivity to reinforcement information ^15,16^. The importance of this construct in depression is now being confirmed in several independent studies ^17-21^.

Here, we looked at changes in depressive patients during treatment with escitalopram, a commonly prescribed antidepressant ^22^ using a novel functional imaging probe of the signal tracking prospective rewards during the effort expenditure phase of reward processing (Figure 1A). Our aim was to investigate the capacity of this probe to identify neural substrates that may track the evolution of depression in patients treated with a serotonergic antidepressant.

**FIGURE 1.**
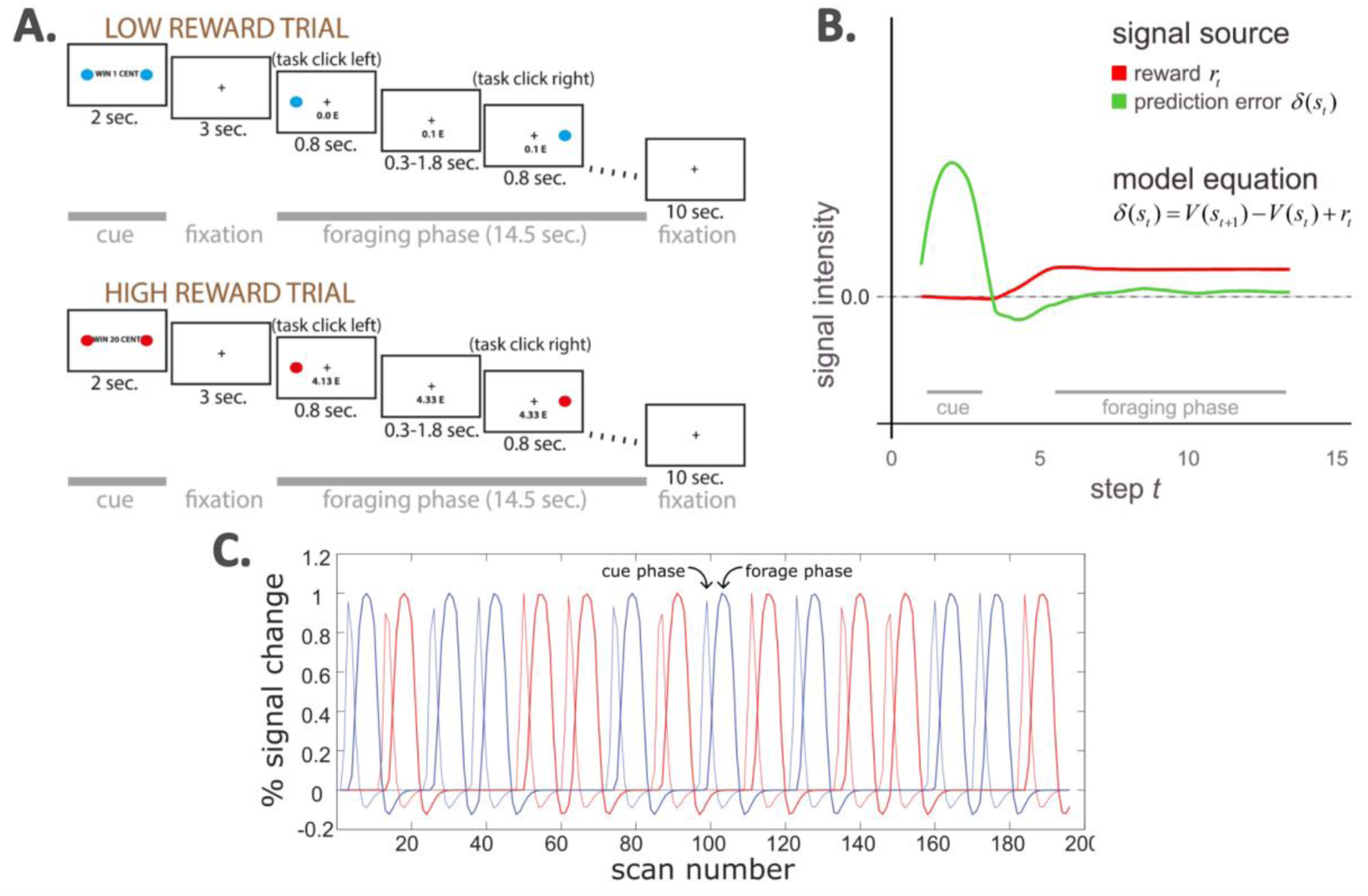
A: *Schematic overview of trial structure in the foraging task.* Participants were first presented with a cue indicating a low- or high-reward trial block, followed by a foraging patch consisting of a rapid sequence of target episodes. Earnings were paid only above a threshold of 20 euros, so that responses were required at both reward levels. B: *Computational model of the cue and forage phases of the task.* At the appearance of the cue, a prediction error computed by a reinforcement learning algorithm signals the availability of reward (green). In the subsequent foraging phase, reward rates (red) are constant but there is no prediction error as the rewards are fully known after the cue. This dissociation allows identifying two distinct motivational drivers when processing reward. C: *BOLD-convolved predictors of the cue and foraging phases.* The timing of the two phases of the task optimizes the number of presentations while maintaining their identifiability (adapted from Orsini et al., 2026 ^23^, CC-BY license).

The design was based on a reinforcement-learning computational model ^24^ (Figure 1B). Each block started with a cue indicating the level of the rewards available during the block (20 or 1 cents). This was followed by targets on the left or the right side of the screen presented at irregular intervals (the ‘foraging’ phase). Participants collected the reward by clicking a button on the side of the target. The cue set the reward level, which remained constant during the block. Earnings were paid out only if a threshold was reached by the end of the experiment, which required participants to respond accurately in both high- and low-reward blocks.

Reinforcement learning captures the neural signal in the reward system arising when new information in the environment signals the availability of reward (‘prediction error’ ^25^; green trace in Figure 1B). In neuroimaging studies, this signal is predominantly visible in the nucleus accumbens (NAcc) ^26^. More recent studies show that a signal tracking reward levels expected from work (‘rewarding context’ ^27^; red trace in Figure 1B) preferentially recruits the ventral tegmental area/substantia nigra (VTA/SN) ^24^. In contrast to prediction error, the rewarding context provides no new information but merely tracks the prospective rewards that may be accrued during work. This quantity is also a key variable of current computational models of foraging ^28,29^. In our study, we mapped the first two phases of reward processing of the psychopathological model described above (the motivational and the intermediate effort expenditure) to prediction error and rewarding context, respectively. In the task, the prediction error was elicited by the cue announcing the reward levels that are available in the subsequent foraging block, while the rewarding context was given by the signal representing the available reward within the block itself^24^ (Figure 1C).

Cues signalling the availability of rewards, i.e. prediction error, have been often linked to motivation in the literature ^30,31^. However, the role of reward representations while working to obtain it has been much less explored. Psychopathological models of effort expenditure have focussed on physical effort and representations of its costs ^17–21^, which in neuroimaging studies modulate activity in the putamen/pallidum ^32^. In our task, in contrast, effort was kept minimal and, through the scoring structure of the task (see Methods), required equally at both reward levels so as to isolate the signal of the rewarding context. This distinguishing feature of our study reflects the working hypothesis that the rewarding context elicits its own distinct signal and may best represent the motivational component of effort expenditure, as is also shown by its association with individual differences in the capacity to pursue long-term goals^23^.

Because this is a naturalistic study without a placebo-controlled patient arm, the design is suited to characterizing the phenotype and its sensitivity to clinical change rather than isolating escitalopram’s pharmacological mechanism. The primary analysis focused on a three-way interaction *reward × group × time*. This interaction captured changes in rewarding context-related processing over time and its modulation by both clinical status and therapy, and was tested in the NAcc and VTA/SN. In a further exploratory analysis we sought evidence for modulation of activity in the dorsal raphe nucleus (DRN), an immediate target of SSRI effects and possible driver of VTA/SN.

By separating the reward context-related signal in the effort expenditure phase from motivational and valuation phases, the study addresses a gap in the literature and offers insight into how this phenotype may represent a clinically important aspect of the depressive syndrome. Such findings may contribute to a better understanding of motivational processes affected by depression and support the development of interventions that help target the full range of reward-processing difficulties.

## 2 Results

### 2.1 Comparison of the Patients and Control Group: Psychopathology

Questionnaire data from the Hamilton Depression Rating Scale (HAMD) and the Snaith– Hamilton Pleasure Scale (SHAPS-D) of the two measurement points were included in the analyses.

For SHAPS-D scores, patients showed a mean change of *M* = −0.46 (*SE* = 0.55, 95% CI [−1.59, 0.66]), which was not statistically significant (*t*(27) = −0.84, *p* = 0.40). Control participants showed a mean change of *M* = −0.42 (*SE* = 0.17, 95% CI [−0.76, −0.07]), which reached statistical significance (*t*(42) = −2.46, *p* = 0.02). Because the control group was untreated and no mechanism would lead healthy participants to become less anhedonic over this interval, we did not interpret this finding further.

For HAMD scores, patients showed a mean change of *M* = −3.50 (*SE* = 1.33, 95% CI [−6.23, −0.77]), indicating a reduction in clinician-rated depressive symptoms across time. This change was statistically significant (*t*(27) = −2.64, *p* = 0.01). In contrast, control participants showed a mean change of *M* = 0.23 (*SE* = 0.42, 95% CI [−0.60, 1.07]), which was not statistically significant (*t*(42) = 0.56, *p* = 0.58). The change in HAMD scores differed significantly between patients and controls (difference = −3.73, 95% CI [−6.57, −0.90]; Welch’s *t*(32.30) = −2.68, *p* = 0.01).

Taken together, these results indicate a significant reduction in clinician-rated depressive symptoms in patients across time, while self-reported anhedonia showed no significant change.

### 2.2 Comparison of the Patients and Control Group: Trial-level Behavioural Results

Linear mixed-effects modelling of reaction time (RT) showed a significant main effect of reward condition, indicating faster responses during high-compared to low-reward trials (*β* = -2.41 ms, 95% CI [-3.73, -1.10], *t* = -3.60, *p* < .001; Figure 2A). A significant main effect of session was also observed (*β* = −3.76 ms, 95% CI [-5.05, -2.47], *t* = −5.73, *p* < 0.001), reflecting faster responses at the second measurement (T2) compared to the first measurement (T1). No significant effect of group (patients vs. control) in reaction times was observed across sessions and reward levels. However, a significant group x session interaction was found (*β* = −4.56 ms, 95% CI [-7.29,-1.83], *t* = −3.27, *p* = .001) indicating faster RT at the second measurement point in patients vs. controls. No significant reward condition x group interaction was observed.

**FIGURE 2.**
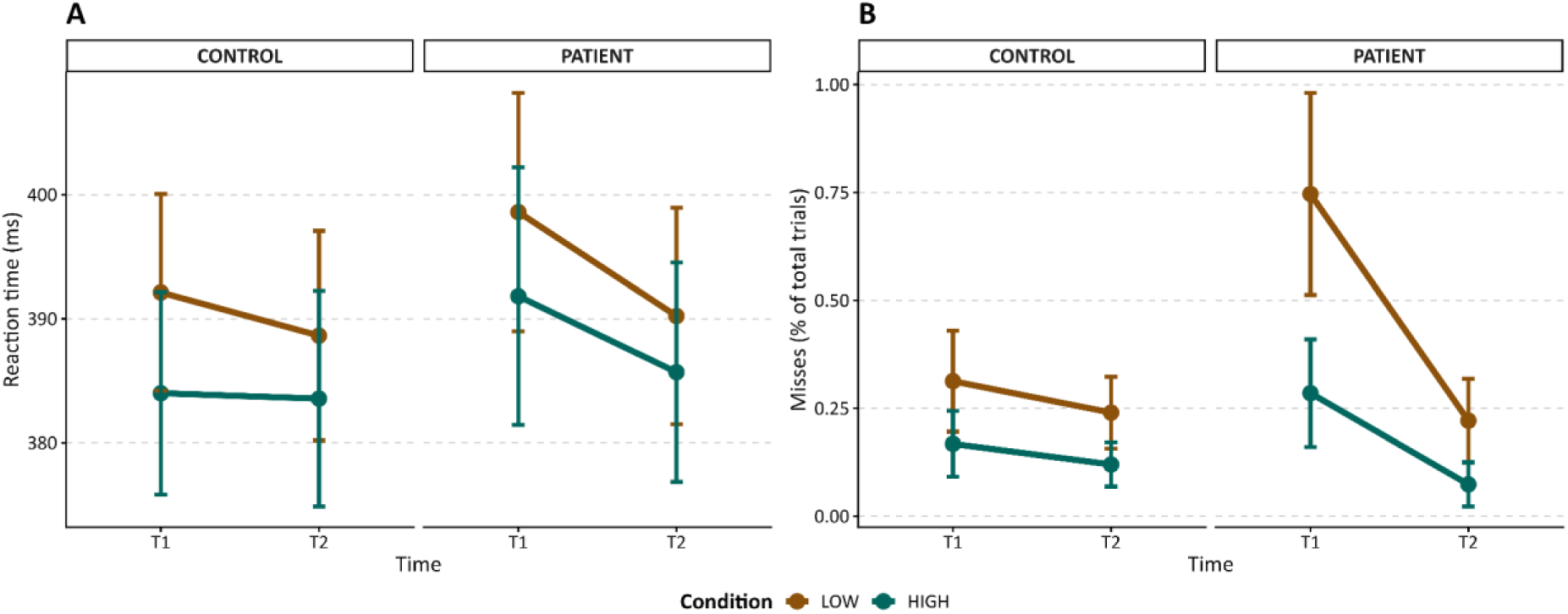
Behavioural results during the foraging task. A: Mean reaction times (ms) as a function of reward condition x time x group. B: Mean missed responses (% of total trials) as a function of reward condition x time x group. Error bars indicate ± SEM.

Generalized linear mixed-effects modelling of response accuracy did not reveal significant effects of reward condition, time, or their interactions when correct responses were analysed (all *p* > .05; see Figure 2B). Because correct responses occurred at ceiling levels, additional analyses examined miss responses. Across all trials, the group × time interaction reached trend level (*p* = .064). Restricting the analysis to the first trial of each block revealed a significant group × time interaction (decrease in odds ratio 0.16, 95% CI [0.04, 0.64], *z* = −2.60, *p* = .009), indicating that patients improved their miss rates in the first trial at T2 relative to T1, whereas performance in the control group remained largely unchanged (our interest in the first trial is due to previous results showing that it is generally handled differently ^33^).

Exploratory analyses further examined whether behavioural performance (RT and accuracy) was associated with VTA activity at T2 while controlling for baseline behavioural performance and baseline VTA activity. No significant associations were observed for either RT or accuracy (all *p* > .05).

### 2.3 Comparison of the Patients and Control Group: Neuroimaging

Before looking at the main contrast of the study, we verified that the high vs. low reward contrast was eliciting the known activations in the cue and foraging phases in the whole sample (collapsing patients and controls and study phases). This was indeed the case (for details, see Figure S3 and Table S1 in the supplements). Specifically, in the foraging phase we obtained robust activity in the VTA.

We then proceeded to test the main hypothesis of the study by conducting a three-way interaction *reward × group × time* contrasting reward magnitude (high vs. low), group (patients vs. control participants), and change over time (second vs. first measurement) separately for the cue phase and the foraging phase. We used the clusters of activation in the whole sample in NAcc and VTA to create region of interest (ROI) analyses focusing on *a priori* hypotheses of involvement of areas associated with reward processing.

#### Cue Phase

During the cue phase, the three-way interaction *(reward × group × time)* did not yield any effects that survived correction for multiple comparisons either in the ROI or the full brain analyses.

#### Foraging Phase

In contrast, during the foraging phase, the three-way interaction *(reward × group × time)* revealed a clear and significant activation pattern. As shown in Figure 3A, patients exhibited increased activity relative to control participants during the second compared with the first measurement, specifically for high versus low reward conditions, in the VTA. The peak activation was located at MNI coordinates *x* = −6, *y* = −16, *z* = −16 (*t* = 2.93). The permutation-based inference revealed that this effect was significant at the cluster level (*k* = 80, *p* = .018, permutation-corrected) and reached peak-level significance (*p* = .021). A complete overview of the neural activation may be found in Table S2. An exploratory whole-brain analysis additionally revealed activity in the right temporal lobe, accompanied by a corresponding effect in the left temporal lobe. However, these effects reached only trend level significance after correction for multiple testing. No effects were observed in the opposite direction, either within the ROI analysis of the mesencephalon or the NAcc or elsewhere in the brain.

**FIGURE 3.**
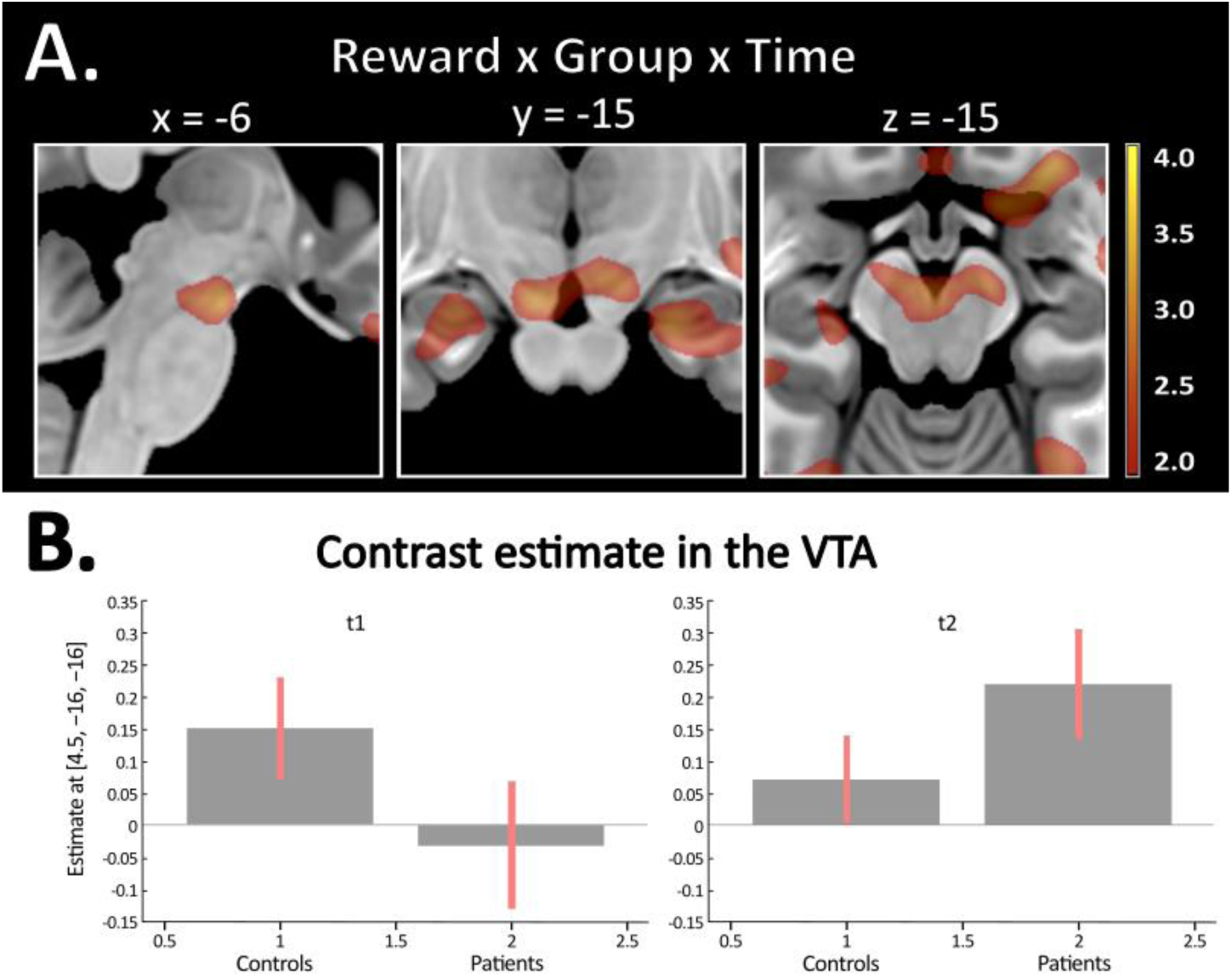
Results of the neuroimaging analyses in the foraging phase. A: Activity in the foraging phase (high vs. low reward; patients vs. controls; second vs. first measurement) on *p* = 0.01 uncorrected. B: Estimation in VTA [-6, -16, -17.5]. The values on the y-axis refer to the percentage change in signal. Left plot: Effects of high vs. low reward in patients and controls at the first measurement time point. Right plot: The same effects at the second measurement time point.

To further clarify the three-way interaction observed during the foraging phase, neural activity was examined separately for patients and control participants at each measurement time point. At the first measurement (T1), parameter estimates suggested higher neural activity in control participants than in patients for the high vs. low reward contrast during the foraging phase (*t* = 2.37; see Figure 3B, left plot). At the second measurement (T2), activity in the control group remained largely unchanged relative to T1 (*t* = 1.18), whereas patients showed an increase in neural activity during the foraging phase (*t* = 3.05; see Figure 3B, right plot). As a result, patients appeared to show higher activity levels than control participants at T2 (*t* = 2.22). However, these apparent group differences at the individual measurement time points did not survive permutation-based correction for multiple comparisons.

#### Cue + Foraging Phase

While the present paradigm was developed to visualize VTA/SN and NAcc activity, we also conducted a separate exploratory analysis to detect activity in the DRN. This activity was absent, even at uncorrected levels, in the previous analysis. The new analysis was conducted with a targeted pipeline, consisting of a smaller smoothing level (FWHM = 4 mm) and a bespoke brainstem denoising model (see Methods for details and Figures S1-2 in the supplementary material). Given the activity profile of DRN in animal studies of reward ^28^, we pooled the cue and foraging phases. Statistical inference was performed within an anatomically defined DRN mask obtained from the Neurobiology Research Unit, Copenhagen University Hospital. As shown in Figure 4A, a significant activation was identified within the DRN at MNI coordinates *x* = −1.5, *y* = −31, *z* = −10 (*t* = 3.35), which survived permutation-based peak-level correction (*p* = .004). At an uncorrected threshold the spatial extent of the effect corresponded closely to the independent anatomical DRN mask, effectively retracing its boundaries (Figure 4A); because the displayed map was not spatially restricted, this anatomical concordance was not imposed by the analysis and is difficult to reconcile with a chance finding, which would not respect the shape of the nucleus. We conducted a post-hoc analysis of the cue and foraging phases separately and observed an effect in both (cue phase, *t* = 3.18, MNI coordinates *x* = −3, *y* = −32, *z* = −10; foraging phase, *t* = 1.96, *x* = 0, *y* = −30, *z* = −8).

**FIGURE 4.**
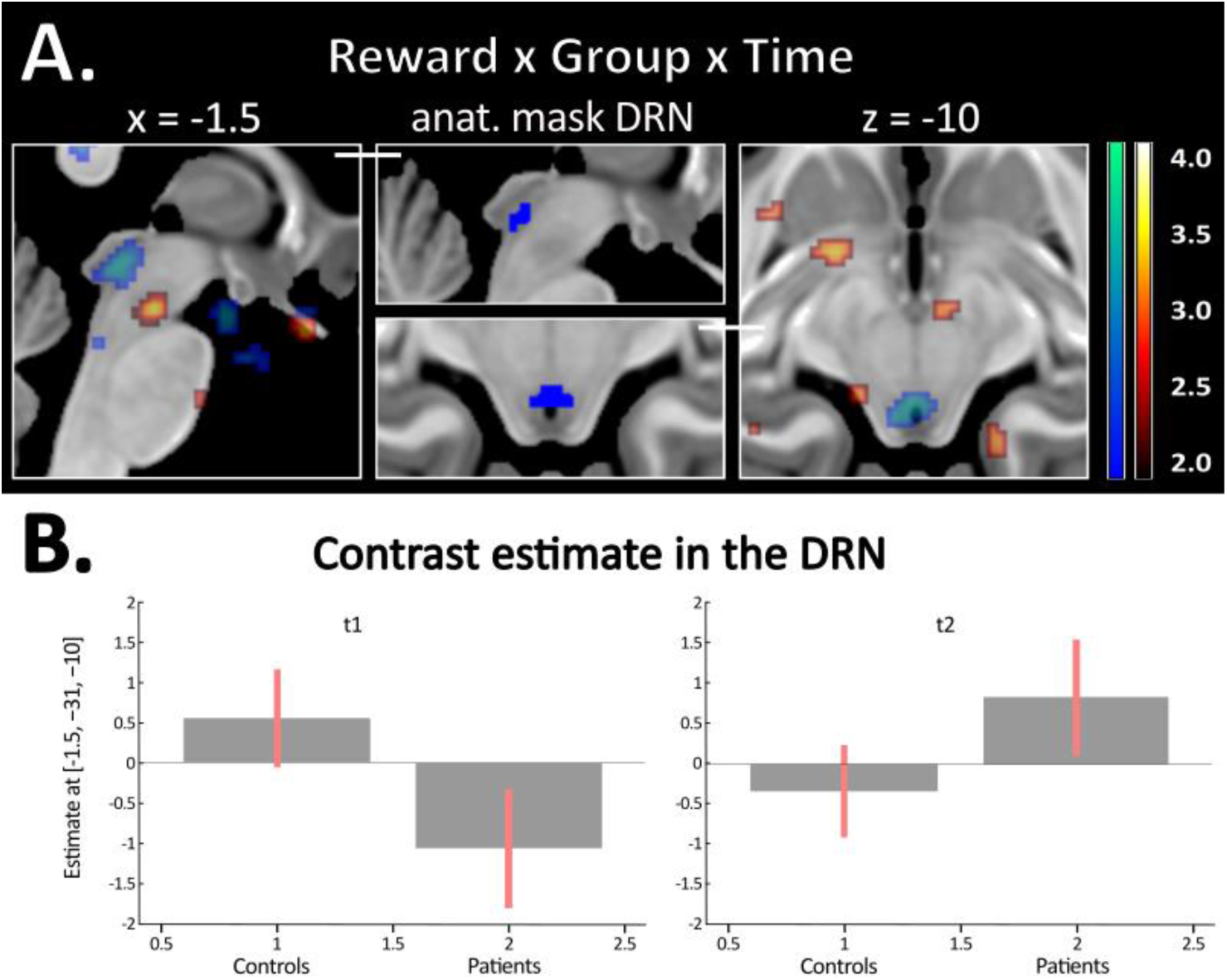
Results of the neuroimaging analyses of the interaction in the pooled cue + foraging phases. A: Contrast activity (high vs. low reward; patients vs. controls; second vs. first measurement) at *p* = 0.01 uncorrected. Red: VTA activity; blue: DRN activity. B: Estimation in DRN [-1.5, -31, -10]. The values on the y-axis refer to the percentage change in signal. Left plot: Effects of high vs. low reward contrast in patients and controls at the first measurement time point. Right plot: The same effects at the second measurement time point.

The corresponding parameter estimates for patients and control participants at both measurement time points are shown in Figure 4B, illustrating the pattern underlying this effect. At T1, patients exhibited lower DRN activity than control participants (*t* = -2.74; see Figure 4B, left plot). At T2, DRN activity increased in patients, whereas activity in the control group remained comparatively stable, inverting the contrast patients vs controls (*t* = 2.10; see Figure 4B, right plot). A complete overview of the neural activation is provided in Table S3. Additionally, we verified that also in this preparation VTA remained active in the forage phase, as in the previous analysis (0, −14.5, −14.5, *t* = 2.62).

#### Task effects, interaction group x time

To exclude a generic effect of medication on BOLD reactivity, we tested the possible effects of escitalopram treatment in patients vs. controls in a contrast pooling both reward levels and phases (cue and forage), i.e. in the effect of the task as a whole relative to the fixation baseline (the strongest signal in the paradigm). This contrast did not reveal any significant effect surviving any level of correction (peak *t* in the whole volume: 3.56, max cluster size, 44 voxels, both *p* > 0.97).

### 2.4 Association with symptom improvement

As reported in Section 2.1, between the first and second measurement there was an improvement in depressive symptoms as measured by the HAMD in patients. To examine whether changes in neural activity were associated with symptom change, an exploratory multiple linear regression analysis was conducted with HAMD scores at the second measurement (T2) as the dependent variable. VTA activity during the reward condition at T2 was entered as the predictor of interest, while VTA activity at T1, baseline HAMD scores at the first measurement (T1), age, and gender were included as covariates. Because VTA activity at T1 is included in the model, the coefficient for VTA activity at T2 reflects the change in VTA activity between sessions. Because the phenotype implies that greater reward-context representation supports motivational function, we specified a priori a directional hypothesis that higher VTA activity would be associated with lower symptom severity, and evaluated it with a one-tailed test.

As shown in Figure 5, higher VTA activity at T2 was associated at trend level with lower HAMD scores at T2 after adjustment for VTA activity at T1, baseline HAMD scores (T1), age and gender (*b* = −9.07, *SE* = 5.51, 90% CI [−18.54, 0.39], *t*(22) = −1.65, *p* = .057, one-tailed). Scanning site and VTA activity at T1 were not influential to demonstrate this effect in the model (model with scanning site, *b* = −9.61, *SE* = 5.68, *t*(61) = −1.69, *p* = 0.053, one-tailed; model without VTA at T1, *b* = −9.25, *SE* = 5.37, 90% CI [-18.45, -0.06], *t*(23) = −1.73, *p* = 0.049, one-tailed). The lack of influence of VTA activity at T1 is due to it being nearly uncorrelated with activity at T2 (*r* = −.02). Activation at T1 did not predict the Hamilton scores at T2 (*b* = 1.21, *p* = .78). The association we observed is therefore not a baseline association carried over to T2. While of moderate size, this effect is directionally consistent with the phenotype and, considered together with the imaging findings, is consistent with the interpretation that greater effort-related neural activity following treatment accompanies lower residual depressive symptom severity.

**FIGURE 5.**
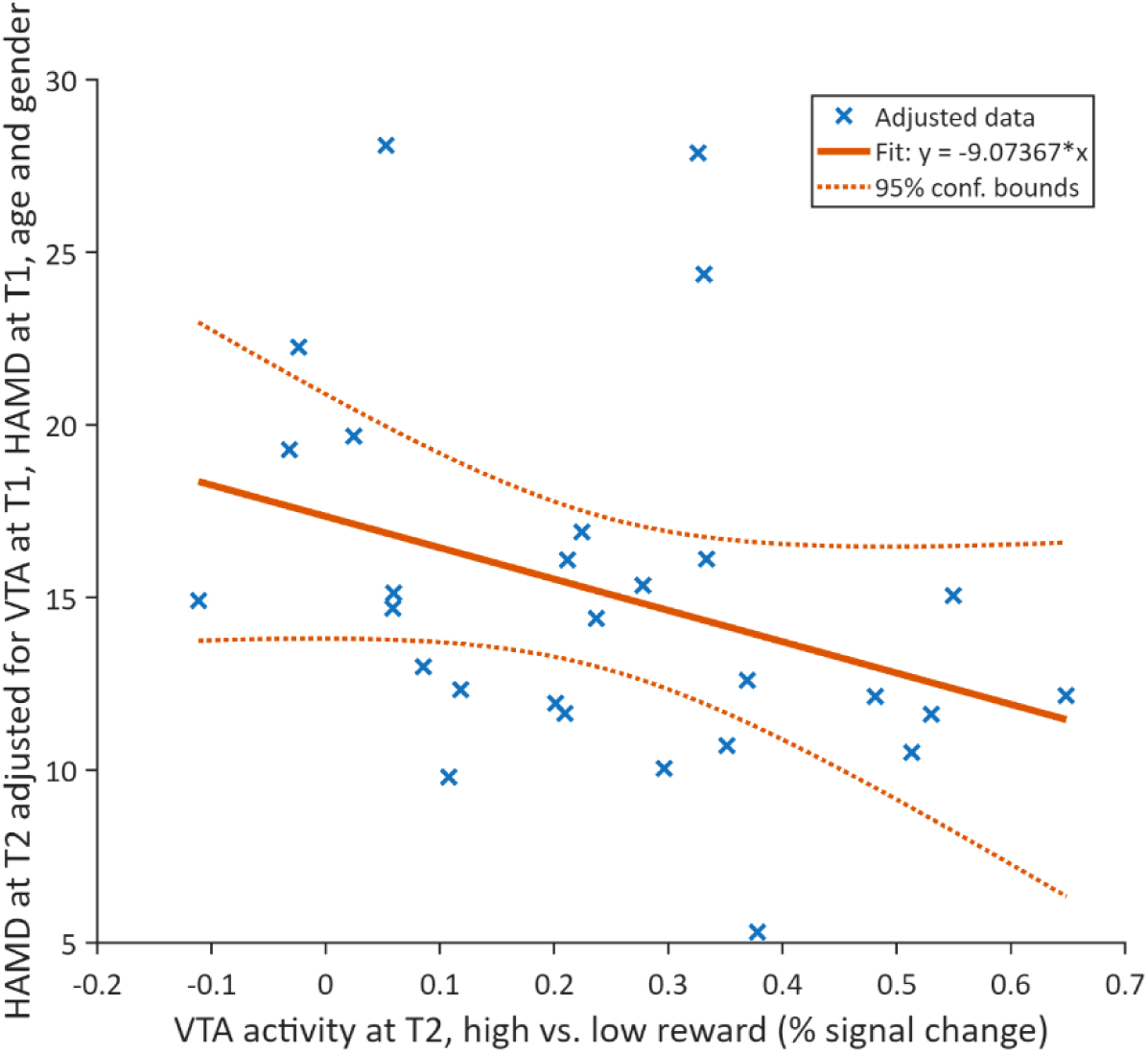
Association between HAMD scores at T2 and VTA activity at T2 after adjusting for VTA activity at T1, baseline HAMD scores (T1), age, and gender. The regression line reflects the partial effect of VTA activity in the multiple regression model.

## 3 Discussion

The present study examined neural activity associated with sustained effort expenditure while working to obtain rewards. Our task was designed to isolate neural processes associated with representations of reward accruing from work (‘rewarding context’, ‘value of work’ ^24,34^) independently of reward anticipation triggered by cues, computation of trade-offs with action costs, and consummatory pleasure. While based on a computational analysis, this approach differs from many existing paradigms in the simplicity of the task, allowing its administration to patient populations without confounding its neural signature with that arising from performance failures and their downstream consequences. This allowed us to address a gap in the human studies on the possible avenues through which changes in depressive symptoms may be accompanied by changes in a key dopaminergic circuit.

### Midbrain reward circuitry in effort expenditure

Reward levels during effort expenditure differentially engaged midbrain reward circuitry, as already shown in previous studies ^24^. This pattern indicates that midbrain reward systems contribute to effort expenditure under delayed reward delivery. This activity is consistent with models of midbrain systems supporting behavioural persistence and sustained engagement when working for rewards over time ^27^. In this respect, the findings provide empirical support for the conceptualization of effort expenditure as a distinct construct within the positive valence system of the RDoC framework ^13^, while isolating the neural substrates associated with the representation of the rewarding context that may support effort expenditure.

Data from this paradigm showed that changes in effort-related activity in the VTA/SN differed between patients and control participants. In VTA/SN, activity increased after treatment in patients. In the exploratory DRN analysis, the reward contrast changed more in patients than in controls. The patient estimate was lower at baseline and higher at follow-up than the corresponding control estimate.

Greater VTA/SN activity after treatment was associated with lower depression severity. Due to lack of randomization and the borderline significance, however, this analysis should be viewed as exploratory and suggestive of the possible utility of this neuroimaging phenotype in future studies. An association between VTA activity and clinical improvement of depression was also recently reported in a randomized controlled trial of biofeedback training ^35^.

Together, these studies provide convergent support on the role of VTA in symptomatic improvement. In our study, although patients also showed improvements in behavioral performance over time, exploratory analyses did not show significant associations between VTA activity and either RT or accuracy. This suggests that the observed neural changes were not directly related to changes in task performance, but instead reflected changes in neural processes supporting sustained effort expenditure. In line with this finding, physical vigour of responding has been located in the putamen/pallidum ^32^.

At the same time, self-reported anhedonia did not change significantly, in line with its treatment resistance as reported in the literature. This finding is consistent with multidimensional models of anhedonia ^2,11,36^ and with clinical observations that serotonergic treatments may alleviate mood symptoms while leaving anhedonia largely unaffected ^9,10^.

This pattern also suggests that the persisting anhedonic deficit may be selective. Recent studies provide evidence that endogenous opioids, and not dopamine, mediate hedonic experience in the consummatory phase ^37,38^.

Although causal inference is limited by the naturalistic design, the observed midbrain neural changes are consistent with a VTA-mediated mechanism through which motivation driving effort expenditure may change with symptomatic improvement.

### Possible role of serotonergic circuits in effort expenditure

Recent research has revised our understanding of serotonin function. Because of the numerous receptor types, the action of dorsal raphe serotonergic neurons defies simple characterization and goes beyond the traditional notion of coding aversive events (for reviews, see Ohmura & Nagayasu, 2025; Koolschijn et al., 2024 ^12,39^). Serotonergic neurons exhibit sustained firing that codes rewarding context ^40^ and promote waiting when rewards are delayed ^41,42^. Unlike typical activity of dopaminergic neurons in NAcc, which fire at the appearance of a cue (Figure 1B, green line), or the preferential activation of VTA/SN during the foraging phase (Figure 1B, red line), dorsal raphe activity in these studies occurs in both phases (resembling the sum of the green and red lines in Figure 1B ^28^).

Serotonergic activity in the dorsal raphe may modulate dopamine function through numerous projections to dopaminergic circuits, including the VTA (for reviews see Nakamura, 2013; Peters et al., 2021 ^40-43^), but the effect of serotonergic activity may depend on the receptor class ^44^. Synergistic function with VTA may be mediated by specialized dual serotonin/glutamate raphe VGluT3-expressing neurons, which directly excite dopamine VTA cells via rapid-acting 5-HT3 receptors that express the serotonin transporter, leading to dopamine release ^45–47^.

These findings have forced a revision of traditional models that viewed serotonin and dopamine function in simple opposition to each other ^48^. Current computational models of the reward-related serotonin signal propose it to emerge from the sustained integration of experienced outcomes to compute the value of prospective rewards ^28^. This integrated signal provides an estimate of the level of the rewarding context and may promote sustained activity ^12,40^. In the foraging study of Lottem et al. (2018)^29^, activity of serotonergic neurons in the dorsal raphe tracked levels of reward in the environment (analogous to reward levels in the present study), and their optogenetic activation increased persistence in working to obtain rewards. In line with this finding, disrupting the dorsal raphe prevented animals from matching their motivation to the reward levels provided by the environment ^49^. As in these studies, DRN activity was present in both the cue and foraging phase. Although outside the scope of our study, DRN activity at the cue would be consistent with the effect of antidepressants on reinforcement learning^50-52^.

However, it should be noted that the projections from the dorsal raphe to VTA are at least of three types (glutamatergic, serotonergic, and dual) and that the excitatory dual type projects to only a subgroup of dopaminergic neurons ^47^, in line with evidence for multiple roles in emotional processing ^8,53^. In contrast to the sustained activity of VGluT3 expressing projections, purely serotonergic neurons appear to stimulate VTA phasically ^54^. Furthermore, much less is known about the relationship between VTA and the median raphe nucleus, even if this latter has been implicated in anxiety ^55^ and in the action of SSRIs ^56^. A further cautionary note is that the fMRI activity registered in the VTA in the foraging phase may not be necessarily dopaminergic, as the action of long-range dual glutamate-GABA VTA neurons would not be inconsistent with the behaviour modelled by this paradigm ^57,58^ (for a discussion, see Orsini et al., 2026 ^33^).

### Clinical and conceptual implications

The identification of an effort-related neural phenotype provides a potential target for future studies. Assessing sustained behavioural engagement may complement existing approaches focused on reward anticipation or choice and may help identify dimensions across which interventions work. Furthermore, while consistent with models in the clinical neurosciences that map motivational processes to dopamine function, our findings also motivatethe hypothesis that traditional views on the lack of capacity of serotonergic medication to support dopaminergic function may have to be revised, a hypothesis that placebo-controlled studies could test.

At a conceptual level, studies of effort expenditure support dimensional models of reward dysfunction and provide empirical grounding for the RDoC framework. Effort expenditure may represent a mechanistic pathway linking disturbances in the reward system to impaired functioning across psychiatric conditions, thereby contributing to a more precise characterization of motivational deficits. The findings support the use of fMRI in the investigation of how serotonin and dopamine interact during sustained reward pursuit and whether this interaction contributes to SSRI response in depression.

### Limitations

The study used a naturalistic design in which patients received escitalopram between measurements, but treatment was not randomized or experimentally controlled. Consequently, causal conclusions regarding serotonergic modulation cannot be drawn. The absence of a placebo or untreated comparison group limits the ability to distinguish medication effects from spontaneous symptom improvement. Nevertheless, our study identified a novel phenotype characterizing motivation in effort expenditure while also showing that DRN activity may be recovered in fMRI data provided a targeted denoising model is used.

An important cautionary note concerns systematic effects of escitalopram administration on the BOLD response, confounded by the naturalistic design with response to medication. SSRIs induce a generalized reduction in baseline cerebral blood perfusion ^59^, which may amplify task-elicited BOLD responses. However, in this case we should have observed a generalized amplification of the task effect in patients at the second timepoint, which was completely absent. Therefore, while we would acknowledge that some of the cortical changes we observed, such as those on the occipital cortical areas, might conceivably be due to such indirect effects, the pattern was inconsistent with a general increase of BOLD responses.

The task was designed to isolate sustained effort expenditure by minimizing decision-making and immediate reward feedback. While this strengthens interpretability, it limits direct comparability with paradigms involving effort-based choice or reward consumption. Anhedonia was assessed using a self-report measure, and subtle changes in hedonic processing may not be captured by subjective ratings. Statistical inference was conducted within an functionally defined mesencephalic ROI; although hypothesis-driven and appropriately controlled, this approach limits conclusions regarding effects outside this region. Finally, sample size and clinical heterogeneity may constrain generalizability.

## 4 Materials and Methods

### 4.1 Participants

#### Recruitment

Patient participants were recruited in the Psychosomatic Clinic (University Clinic Bonn), the Psychiatry and Psychotherapy Clinic (University Clinic Ulm), and the Psychosomatic Clinic (also University Clinic Ulm). Patients were informed about the study by their treating physician, who also checked the inclusion and exclusion criteria. Control participants were contacted through local announcements.

#### Inclusion criteria

Participants were between 18 and 65 years of age, right-handed, and had normal or corrected-to-normal vision. All had a body mass index (BMI) above 18, were in good general health, and showed no structural brain abnormalities. All participants were Caucasian and participation was voluntary. Depressed patients (DP) were required to meet criteria for a current depressive episode within the context of a unipolar affective disorder (ICD-10: depressive episode F32, recurrent depressive disorder F33, dysthymia F34), diagnosed by the treating physician and confirmed with the Mini International Neuropsychiatric Interview (MINI), which was also used to rule out other psychiatric disorders. Patients had a Hamilton Depression Rating Scale (HAMD) score of at least eight, and must not have taken SSRIs or other antidepressant medication within the preceding two months. None of the patients had received any treatment for depression before the study.

#### Exclusion criteria

Participants were excluded if they were pregnant, reported regular intake of medication (in patients, L-thyroxine, oral contraceptives, statins and ACE inhibitors were permitted), had taken anticoagulants, nonsteroidal anti-inflammatory drugs or beta blockers in two weeks before the study, had a diagnosis of a psychotic, bipolar or schizoaffective disorder, substance dependence or a relevant somatic illness, or were currently suicidal.

Standard contraindications for MRI scanning also constituted exclusion criteria ^60^. A total of *N* = 75 participants were pre-screened for the study; two were excluded because they did not meet inclusion criteria. Two additional participants were excluded from further analyses due to incomplete task performance during MRI scanning (see below, Data preprocessing).

#### Final sample

The final sample (*N* = 71) included 28 outpatients with a current depressive episode without psychotic symptoms (15 female, *M* = 36.18 years, *SD* = 12.72, age range 19-63 years) and 43 healthy control subjects (20 female, *M* = 33.44 years, *SD* = 10.74, age range 19–56 years). Patients and controls did not differ in age (*t*(69) = 0.98, *p* = .33) or sex (χ²(1) = 0.34, *p* = .56). At baseline, patients’ HAMD scores averaged 18.64 (*SD* = 5.29, range 8–33) and their SHAPS-D scores 3.46 (*SD* = 2.63, range 0–9). All participants were Caucasian, native German speakers and right-handed. Participants with corrected-to-normal vision wore MRI-compatible glasses.

#### Sites

Participants were scanned in Bonn at the German Center for Neurodegenerative Diseases (19 patients, 27 controls) and Ulm at the Department of Psychiatry and Psychotherapy III (9 patients, 16 controls), and each participant was scanned on the same scanner at both sessions. The proportion of patients did not differ between sites (χ²(1) = 0.19, *p* = .66), and neither did age (*t*(69) = −0.62, *p* = .54) or sex (χ²(1) = 0.11, *p* = .74). Among patients, baseline HAMD scores did not differ significantly between sites after adjusting for age and sex (Bonn vs. Ulm: *b* = 3.26, 95% CI [−0.90, 7.43], *p* = .12), nor did HAMD scores at T2 after additionally adjusting for baseline HAMD (*b* = −0.48, 95% CI [−5.62, 4.66], *p* = .85).

### 4.2 Study Design

#### 4.2.1 Procedure

Participants were initially screened for suitability via email or telephone. This screening included a review of the inclusion and exclusion criteria as well as a detailed explanation of the study objectives, the experimental procedure, and potential side effects associated with MRI scanning. Written informed consent was obtained from all participants prior to participation.

Before MRI scanning, the experimental paradigms were explained and practiced on a computer in the preparation room, allowing participants to familiarize themselves with the foraging task. The MRI session lasted 50 minutes. After scanning, participants completed questionnaires assessing demographic variables, past experiences, and current emotional state.

Immediately after the first scan, patients started escitalopram at 10 mg per day. Apart from escitalopram, patients received no pharmacological or psychotherapeutic treatment between the two sessions, and none was hospitalized. A second MRI session was conducted three weeks later (21–26 days after the first scan). The same experimental paradigms were used, and the questionnaires were administered again. This interval was chosen to allow antidepressants to unfold their effect. Classic neurophysiological studies in the mouse have established that the administration of SSRIs causes a compensatory drop in the firing rate of neurons in the dorsal raphe, which gradually recovers in 2-3 weeks after the habituation of the 5-HT1A autoreceptor ^61^ (for a recent perspective, see Henningson et al., 2026 ^62^). Therefore, at three weeks of treatment with escitalopram we should expect DRN activity to have recovered from the initial compensatory feedback loop.

#### 4.2.2 Foraging Task

The task consisted of a cue and of continuous sustained attention “foraging” phase (Figure 1A ^24^). In the *cue phase*, participants were informed about the reward magnitude (low vs. high) associated with the upcoming trial block. This phase reflected changes in reward expectation (motivational phase) and did not require any behavioural response. In the subsequent *foraging phase*, participants performed rapid responses to visual stimuli over an extended sequence of trials, requiring continuous attention and sustained effort. Importantly, rewards were not delivered immediately but paid out only after a delay of one month. This design minimized the influence of prediction error/incentive salience processes and immediate hedonic responses, isolating neural activity associated with sustained effort expenditure ^24,27^ (see Figure 1B).

The task consisted of 18 blocks. A fixation cross was continuously displayed at the centre of the screen. At the beginning of each block, the reward magnitude for correct responses was announced (*cue phase*) and displayed for 2 seconds. Reward magnitude was either low (1 cent) or high (20 cents) and remained constant within each block. Following the cue phase, the fixation cross was presented alone for 3 seconds. After this, 12 targets were presented sequentially, each for 0.8 seconds separated by intervals of varying randomly according to an exponential schedule bounded between 300 and 1800 msec, with an average interval of about 1220 msec, either to the left or right of the fixation cross at random (foraging phase). Participants were instructed to press the left or right button on the response console as quickly as possible, depending on the dot location. Between the end of the foraging phase and the next cue the fixation cross was displayed alone for 10 sec.

For each correct response, participants earned the announced reward amount, which was accumulated in a virtual currency. The total accumulated amount was continuously updated and displayed. At the end of the experiment, the virtual earnings were converted into real money only if participants had reached a minimum threshold of 20 units (i.e., 20 euros). Participants were also informed that the reward would be transferred to their bank account one month later.

The sequence of reward conditions was arranged such that this threshold could only be reached toward the end of the task. Participants were informed at the outset that they could not afford many errors in order to reach the threshold, which encouraged them to maintain attention and respond accurately throughout both low- and high-reward blocks. This design required participants to sustain effort throughout the entire task.

#### 4.2.3 Questionnaires

##### Demographic data

Participants provided demographic information, including age, gender, origin, and social characteristics such as occupation, marital status, income, and socioeconomic status.

##### Hamilton Depression Rating Scale (HAMD)

The severity of depressive symptoms was additionally assessed using the Hamilton Depression Rating Scale (HAMD) ^63^. The HAMD is a clinician-rated scale that evaluates depressive symptoms experienced during the past seven days, including depressed mood, guilt, loss of interest, suicidal ideation, sleep disturbances, psychomotor retardation or agitation, somatic symptoms, anxiety, and related features. The scale consists of 21 items rated on either a three-point scale (0–2) or a five-point scale (0–4). In the present study, the 21-item German version ^64^ was used. Previous reviews report high internal consistency for the total HAMD score, with Cronbach’s alpha values ranging from .46 to .97, as well as good inter-rater reliability (*r* = .82–.98) and test–retest reliability (*r* = .81–.98) ^65^. Convergent validity has been demonstrated through correlations with the Brief Psychiatric Rating Scale ^66^, ranging from *r* = .56 to *r* = .89. Studies of the German version report satisfactory internal consistency across depressive samples (α = .73–.85) ^67^.

##### Snaith-Hamilton Pleasure Scale (SHAPS-D)

Anhedonia was assessed using the German version of the Snaith–Hamilton Pleasure Scale (SHAPS-D), a self-report measure designed to capture subjective anhedonia in psychiatric populations. The questionnaire assesses interest and pleasure across domains, including social interaction, sensory experience, and eating behaviour. Previous studies have demonstrated the scale’s good reliability and validity ^e.g., 68^. Item scores are summed to yield a total score, with higher values indicating greater levels of anhedonia.

### 4.3 fMRI Data Acquisition

Participants were scanned at the Department of Psychiatry and Psychotherapy III of the University of Ulm and at the German Center for Neurodegenerative Diseases (DZNE) in Bonn. At these sites, functional MRI data were acquired using a 3-Tesla Magnetom Prisma scanner (Siemens, Erlangen, Germany) or a 3-Tesla Magnetom Skyra scanner (Siemens, Erlangen, Germany), both equipped with a 64-channel head coil. During scanning, participants lay supine in the scanner, held a response console, and viewed the foraging task on a projection screen via a mirror mounted on the head coil.

Participants spent approximately three hours at the clinic on the day of the examination, including 50 minutes in the MRI scanner. Functional images were acquired using a T2*-weighted echo-planar imaging (EPI) sequence with the following parameters: repetition time (TR) = 2460 ms, echo time (TE) = 30 ms, field of view (FOV) = 19.2 cm, flip angle (FA) = 82°, matrix size = 64 × 64, voxel size = 3 × 3 × 3 mm (slice thickness 2.5 mm with a 0.5 mm gap), and 39 slices.

### 4.4 Neuroimaging Analysis

#### 4.4.1 Data Preprocessing

Data preprocessing, statistical modelling, and correction for multiple comparisons were performed using Statistical Parametric Mapping (SPM12; Wellcome Trust Centre for Neuroimaging, London, UK; www.fil.ion.ucl.ac.uk/spm/) implemented in MATLAB R2013b (version 8.2.0.701; The MathWorks Inc., Natick, USA). Imaging data were realigned using MATLAB (The MathWorks Inc., R2018a, USA) and spatially normalised to the Montreal Neurological Institute (MNI) template based on the Automated Anatomical Labeling Atlas ^69^. Spatial smoothing was applied using Statistical Parametric Mapping (SPM) with a full-width at half-maximum (FWHM) Gaussian kernel of 8 mm.

The analysis targeting the dorsal raphe nucleus was conducted with a smoothing kernel of FWHM 4mm. Denoising for the brainstem used the CompCor technique ^70^. Confounder covariates were added to the first level model for the mean signal from the 4th ventricle (identified by an anatomical ROI), two additional principal components, and their Volterra kernel expansions (first derivative and quadratic term). Mean and 2 principal components from a ROI mask for the internal carotid were added, obtained from a vascular density atlas available online (https://neurovault.org/collections/1061/) ^71^, giving a total of 12 denoising confounder terms. The signal for the confounding covariate terms was extracted from non-smoothed volumes. Effects of denoising on residual mean squares and signal to noise ratios are in the Supplemental information (Figures S1-S2).

Trials in which participants failed to respond were modelled separately as a perturbation regressor. Participants who failed to respond or did not press the response console more than 12 times in total were excluded from further analysis (two participants).

Spatial registration included a segmentation step incorporating spatial priors for the pallidum, red nucleus, and the iron-rich region of the substantia nigra ^24^. Trials were divided into cue and foraging phases and modelled separately under low- and high-reward conditions.

#### 4.4.2 Statistical Analyses

##### 4.4.2.1 Trial-level Behavioural Analyses

Statistical analyses of behavioural data were conducted in R (R Foundation for Statistical Computing, Vienna, Austria) using linear mixed-effect models (LMMs) and generalized linear mixed-effects models (GLMMs) implemented in the *lme4* package. Separate analyses at trial level were conducted for RT (ms) and accuracy (%).

Reaction times were analysed using LMMs with reward condition (high vs. low reward), session (T1 vs. T2), and group (patients vs. healthy controls) as fixed effects, including all interaction terms. To account for task-related variation, first trial within each block, block number, interstimulus interval (ISI), and performance index (PI) were included as additional fixed effects. Participant-specific intercepts were modelled as random effects. Statistical significance was assessed using Satterthwaite approximations of the denominator degrees of freedom implemented in the *lmerTest* package.

Response accuracy was analysed using GLMMs with a binomial error distribution. Separate models were fitted for correct responses and miss responses. Fixed effects included reward condition, time, group, and their interactions. Participant-specific intercepts were included as random effects. Because miss responses occurred infrequently, a simplified model was used to ensure model convergence. An additional exploratory analysis examined miss responses restricted to the first trial of each block.

To examine whether behavioural performance was associated with neural activity, exploratory linear regression analyses related behavioural measures at the second measurement to VTA activity at T2 while controlling for baseline behavioural performance and baseline VTA activity.

##### 4.4.2.2 Neuroimaging Analyses

Statistical models were estimated separately for each participant at the first level. The phases of the task blocks were separately convolved with a canonical BOLD function to produce the predictors at the first level of the analysis (Figure 1C). Movement parameters were included, and volumes with movements exceeding 2 mm were excluded from the analysis. At the second level, contrast estimates were tested with a one-sample *t* test with site, age and sex included as additional covariates.

In a separate analysis, the relationship between depressive symptom severity and neural activity at the second measurement was examined using a multiple linear regression model. HAMD scores at T2 were specified as the dependent variable and VTA activity at T2 was entered as the predictor of interest. VTA activity at T1, baseline HAMD scores at T1, age, and sex were included as covariates. This model assessed whether effort-related neural activity at T2 explained variance in depressive symptom severity beyond that accounted for by baseline symptom severity and demographic factors.

Statistical inference was performed within a mesencephalic region of interest (ROI) based on a priori hypotheses regarding the involvement of midbrain reward circuitry. To define this ROI, the main effect of reward magnitude (high vs. low reward) during the foraging phase was estimated across all participants and both measurement sessions. The resulting activation within the anatomical boundaries of the mesencephalon was thresholded at *p*_uncorr_ < .000001 to define the mask used for the subsequent ROI-based inference ^72^. In addition, exploratory whole-brain analyses were performed; effects identified at the whole-brain level did not survive correction for multiple comparisons and are therefore reported only at a trend level for future meta-analyses.

To obtain robust control of the family-wise error rate, significance testing was based on non-parametric permutation inference. Statistical significance was assessed using 8000 permutation resamples, in which the regressor of interest was randomly permuted across participants. For each permutation, the maximal *t*-values across voxels (for peak-level inference) and the size of the largest suprathreshold cluster (for cluster-level inference) were recorded.

For visualization in figures, parametric statistical maps are reported at an uncorrected threshold (*p* = .01) but were not used for statistical inference.

## Data Availability

All data produced in the present study are available upon reasonable request to the authors after verification that the purposes for any new analyses are consistent with those to which the participants agreed in their informed consent.

## 5 Additional Information

### Ethics approval

The present study was part of a larger project entitled “Investigation of the long-lasting inhibition of the serotonin transporter in the human brain via functional imaging (SERT)”. SERT was approved by the Ethics Committee of the University of Ulm (reference number: 290/14 - Zo/Sta) and the Ethics Review Committee of the University of Bonn (file number: 186/13). The SERT project was conducted and evaluated in accordance with the Declaration of Helsinki, including its current revisions ^73^, the German Medicines Act, the GCP guidelines (Guideline for Good Clinical Practice E6), and the regulations for good scientific practice.

### Informed consent

All participants gave written informed consent before enrolling in the study.

### Compensation

All participants received €50 immediately after the scan. Each participant also received a CD containing structural images of their brain. In addition, the €20 sum that could be won by reaching the threshold in the task was paid out approximately one month after the second fMRI scanning session.

## Supplemental Information

### Reinforcement learning model

Blocks were modelled as a sequence of discrete states (pre-cue, cue, interval, and target/non-target episodes in the foraging block), a finite-state approximation to continuous time. High- and low-reward blocks occurred in random order. State values *V*(*s*_*t*_) = *E*[*r*_*t*_ + *γr*_*t*+1_+. . . ], with *t* indexing steps within a block (the foraging phase) and *r*_*t*_ the reward delivered at step *t* (one target presentation), were learned with undiscounted TD(0) (*γ* = 1) by updating *V*(*s*_*t*_) ← *V*(*s*_*t*_) + *αδ*(*s*_*t*_), where

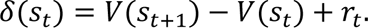

After training, the cue produces a prediction error, positive for high and negative for low reward relative to the pre-cue expectation, because its reward level is unpredictable. In the foraging block, expectations match rewards, *δ* ≈ 0 on average, and the ‘consistency condition’ *r*_*t*_ = *V*(*s*_*t*_) − *V*(*s*_*t*+1_) holds. Smoothed over time, *r*_*t*_ gives the block mean reward rate, interpretable as a perceived reward opportunity. The model therefore predicts that making cues predictable abolishes the cue signal but not the reward-rate signal (tested in Viviani et al. 2020, Experiment 2), and that the foraging block signal does not depend on irregular target timing (tested in Experiment 3). We ran the trained model for 200 trials and plotted smoothed, trial-averaged δ and *r*_*t*_ to reflect the low-frequency BOLD signal generated by these values. More details and a theoretical discussion are in Viviani et al. (2020)^1^. The model was not fitted to the data: it motivates the separation of cue and foraging phases in the design, which were modelled with categorical predictors (Section 4.4.2.2).

Note that the experiment in the scanner does not model the training process; the simulations show how the model represents rewarding context and prediction error after training: participants in the scanner had been informed of the structure of the task and had practiced it before going into the scanner. In both cases, it is the post-training signals that are investigated.

The purpose of the model is to show that, after reinforcement learning, there are no prediction errors concerning the magnitude of reward in the forage block phase, i.e. when participants press the button and collect the rewards; instead, there is a representation of the reward accruing from doing the work (if the reward stopped, there would be a negative prediction error, so the incoming reward must be represented internally by the model to compute a comparison). Our task is set up to investigate the location of this representation, a form of reward processing but one that differs from prediction error.

### Effects of denoising

**Figure S1.**
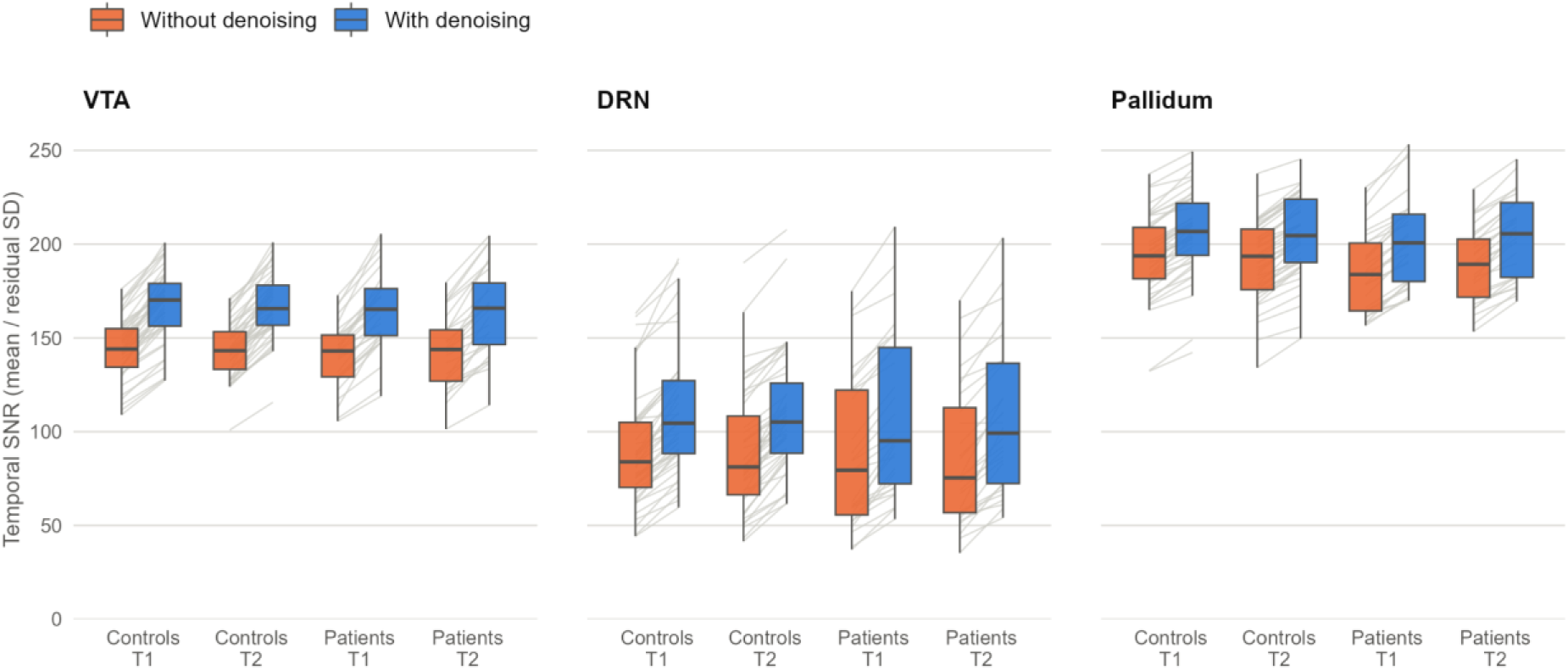
Temporal signal-to-noise ratio (tSNR) in the VTA/SN, the DRN, and the pallidum, without (orange) and with (blue) denoising, by group and session. tSNR was computed at each voxel as the mean of the time series (data smoothed at FWHM 4 mm, scaled to a global mean of 100) divided by the residual standard deviation of the first-level model, computed separately for each session, and averaged over the voxels of each region. Lines join the values of each participant and session in the two models. Boxes show medians and interquartile ranges; whiskers extend to 1.5 times the interquartile range.

**Figure S2.**
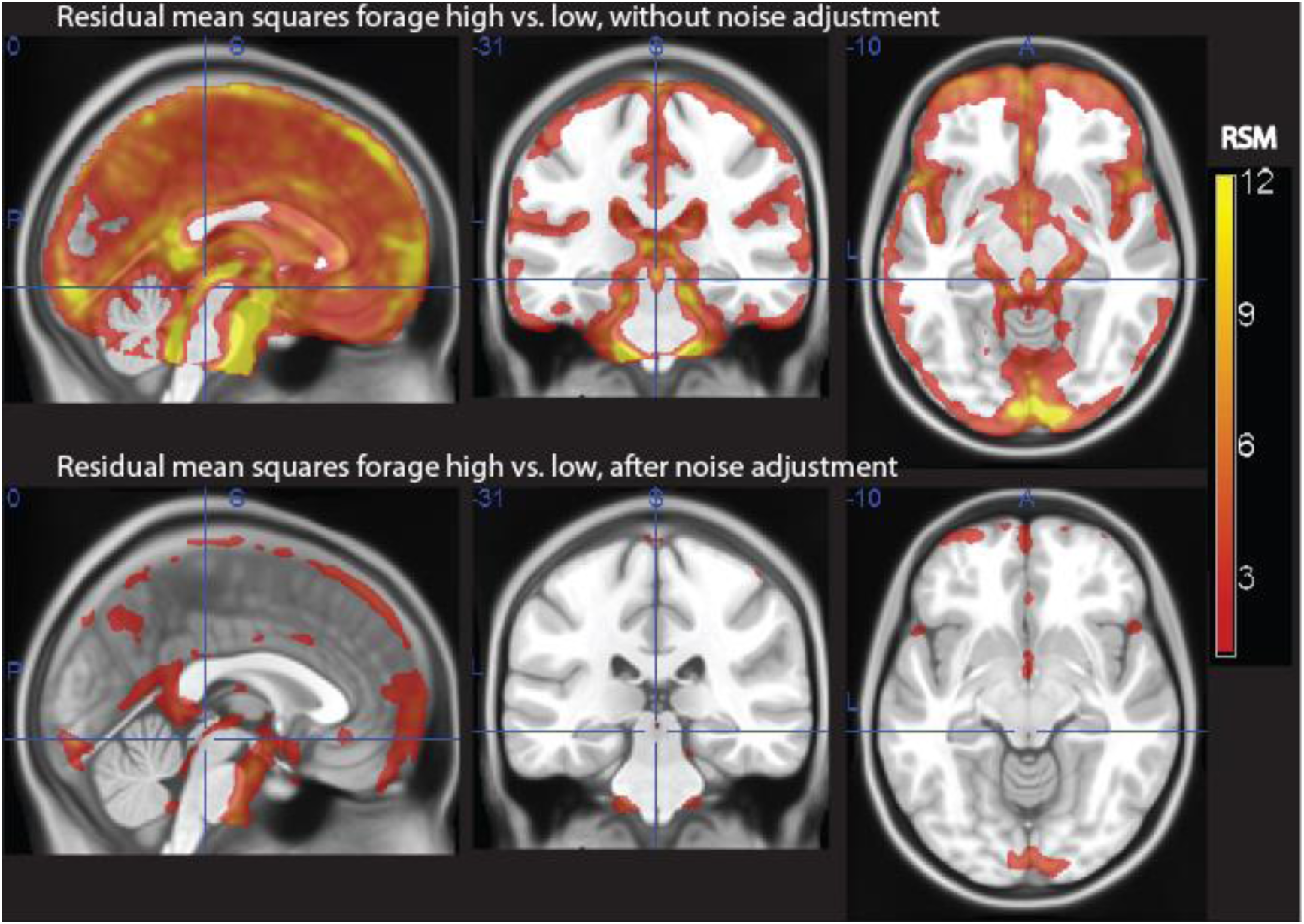
Residual means squares in the forage high vs. low contrast, without (top) and with adjustment (bottom). Smoothing FWHM 4mm. The crosshair is at the peak DRN effect. Data thresholded at 1.2 square percent change.

### Activity in the Reward Condition

The main effect of reward magnitude (high vs. low reward) was examined separately for the cue phase and the foraging phase across all participants (*N* = 71), combining patients and control participants across both measurement time points. Figure S1 illustrates representative activation patterns for both task phases. The displayed coordinates correspond to the mesencephalic regions identified in the main interaction analyses and are shown for an easier comparison with the primary findings reported in the main text. A complete overview of significant activations is provided in Table S1.

#### Cue Phase

During the cue phase, the contrast of high versus low reward revealed robust activation across a distributed reward-related and attentional network (Figure S3A). The strongest activation was observed in a large frontoparietal cluster with a peak at MNI coordinates *x* = −34.5, *y* = −7, *z* = 62 (*t* = 7.65, *p*_FWE-corr_ < .001). Additional significant activation was identified in ventral striatal regions associated with reward anticipation (*x* = 18, *y* = −16, *z* = 21.5; *t* = 6.96, *p*_FWE-corr_ < .001) ^1^, as well as in occipital visual regions (*x* = 37.5, *y* = −80.5, *z* = −10; *t* = 6.49, *p*_FWE-corr_ < .001). The remaining significant activations are reported in Table S1.

#### Foraging Phase

As illustrated in Figure S3B, the analysis revealed significant activation in the VTA and SN, with a peak located at *x* = 3, *y* = −16, *z* = −15 (*t* = 6.36; *p*_uncorr_ < .001; *p*_FWE-corr_ < .001). Correction for multiple comparisons was performed using an anatomical mask of the mesencephalon.

During the foraging phase, the contrast of high versus low reward revealed significant activation in midbrain and cortical regions associated with sustained task performance (Figure S1B). The strongest activation was observed in a large occipitotemporal cluster with a peak at MNI coordinates x = 42, y = −59.5, z = −13 (t = 9.79, *p*_FWE-corr_ < .001).

Additional activation was observed in mesencephalic regions encompassing the ventral tegmental area (VTA) and substantia nigra (SN) (x = −30, y = 17, z = −8.5; t = 9.25, *p*_FWE-corr_ < .001), as well as in frontal cortical regions (x = 42, y = 6.5, z = 29; t = 8.73, *p*_FWE-corr_ < .001). Additional significant activations are reported in Table S1.

**FIGURE S3.**
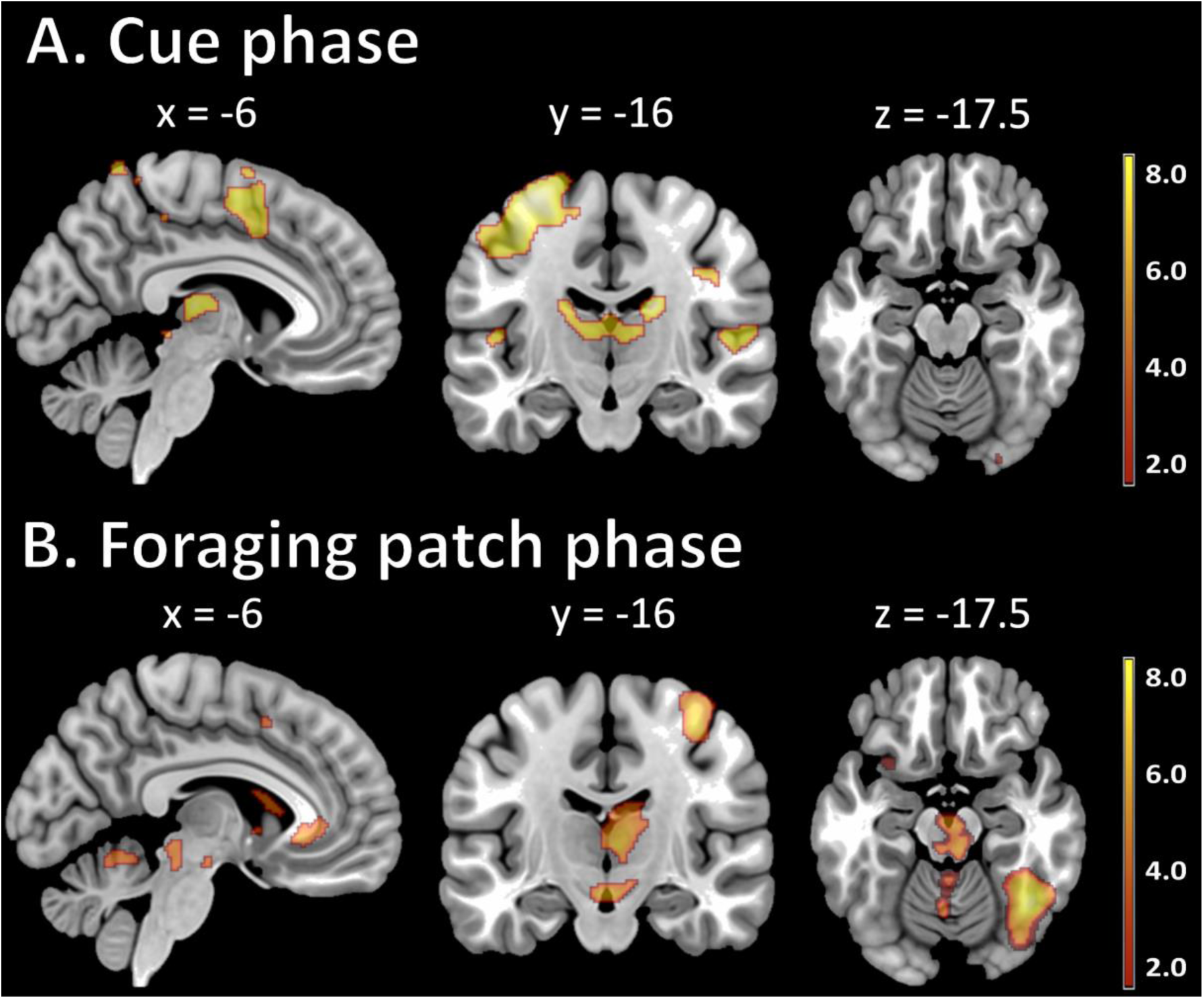
Neural activation during the cue and foraging patch phases. **A:** Activation in the cue phase (high vs. low reward; first and second measurement) across all participants (FWE-corrected). **B:** Activation in the foraging phase (high vs. low reward; first and second measurement) across all participants (FWE-corrected).

**Supplementary Table S1.** Cue and foraging patch phases.

| CUE PHASE |  |  |  |  |  |  |  |  |  |  |
|---|---|---|---|---|---|---|---|---|---|---|
| set |  | cluster |  |  | peak |  |  | MNI coordinates |  |  |
| p | c | p<sub>FWE-corr</sub> | k<sub>E</sub> | p<sub>uncorr</sub> | p<sub>FWE-corr</sub> | T | p<sub>uncorr</sub> | x | y | z |
| 0.00 | 35 | 0.00 | 9007 | 0.00 | 0.00 | 7.65 | 0.00 | -34.5 | -7 | 62 |
|  |  | 0.00 | 3048 | 0.00 | 0.00 | 6.96 | 0.00 | 18 | -16 | 21.5 |
|  |  | 0.00 | 373 | 0.00 | 0.00 | 6.94 | 0.00 | 13.5 | 57.5 | -8.5 |
|  |  | 0.00 | 467 | 0.00 | 0.00 | 6.49 | 0.00 | 37.5 | -80.5 | -10 |
|  |  | 0.00 | 583 | 0.00 | 0.00 | 6.14 | 0.00 | -30 | 39.5 | 35 |
|  |  | 0.00 | 344 | 0.00 | 0.00 | 6.10 | 0.00 | 34.5 | -2.5 | 56 |
|  |  | 0.00 | 342 | 0.00 | 0.00 | 6.06 | 0.00 | -15 | 44 | 0.5 |
|  |  | 0.00 | 81 | 0.06 | 0.00 | 5.93 | 0.00 | -15 | -25 | -10 |
|  |  | 0.00 | 646 | 0.00 | 0.00 | 5.90 | 0.00 | 46.5 | -11.5 | 29 |
|  |  | 0.00 | 326 | 0.00 | 0.00 | 5.90 | 0.00 | 51 | -16 | 6.5 |
|  |  | 0.00 | 147 | 0.01 | 0.00 | 5.79 | 0.00 | -46.5 | -17.5 | 8 |
|  |  | 0.00 | 152 | 0.01 | 0.00 | 5.77 | 0.00 | -12 | -92.5 | -4 |
|  |  | 0.00 | 145 | 0.01 | 0.01 | 5.63 | 0.00 | -18 | 15.5 | 15.5 |
|  |  | 0.00 | 61 | 0.09 | 0.01 | 5.62 | 0.00 | 25.5 | -23.5 | 72.5 |
|  |  | 0.00 | 80 | 0.06 | 0.01 | 5.58 | 0.00 | 18 | 45.5 | 14 |
|  |  | 0.00 | 72 | 0.07 | 0.01 | 5.53 | 0.00 | 27 | -91 | 5 |
|  |  | 0.00 | 69 | 0.07 | 0.01 | 5.49 | 0.00 | -19.5 | 26 | -2.5 |
|  |  | 0.00 | 104 | 0.03 | 0.01 | 5.47 | 0.00 | 24 | -43 | 72.5 |
|  |  | 0.01 | 40 | 0.16 | 0.01 | 5.41 | 0.00 | 16.5 | -11.5 | 60.5 |
|  |  | 0.01 | 38 | 0.17 | 0.02 | 5.39 | 0.00 | -21 | -7 | -8.5 |
|  |  | 0.02 | 17 | 0.36 | 0.02 | 5.37 | 0.00 | -9 | 8 | -2.5 |
|  |  | 0.00 | 111 | 0.03 | 0.02 | 5.34 | 0.00 | -3 | -46 | 63.5 |
|  |  | 0.01 | 24 | 0.28 | 0.02 | 5.29 | 0.00 | 7.5 | 26 | 20 |
|  |  | 0.02 | 17 | 0.36 | 0.02 | 5.29 | 0.00 | -28.5 | 53 | 24.5 |
|  |  | 0.02 | 10 | 0.49 | 0.02 | 5.28 | 0.00 | -9 | 9.5 | 15.5 |
|  |  | 0.02 | 16 | 0.37 | 0.02 | 5.26 | 0.00 | -4.5 | -34 | -4 |
|  |  | 0.00 | 61 | 0.09 | 0.03 | 5.22 | 0.00 | 12 | -44.5 | 54.5 |
|  |  | 0.03 | 8 | 0.54 | 0.04 | 5.13 | 0.00 | -28.5 | 32 | 2 |
|  |  | 0.02 | 15 | 0.39 | 0.04 | 5.13 | 0.00 | -39 | 15.5 | 0.5 |
|  |  | 0.03 | 7 | 0.57 | 0.04 | 5.10 | 0.00 | 55.5 | 3.5 | 0.5 |
|  |  | 0.03 | 4 | 0.68 | 0.04 | 5.10 | 0.00 | 37.5 | 8 | 9.5 |
|  |  | 0.03 | 8 | 0.54 | 0.04 | 5.09 | 0.00 | -7.5 | -35.5 | 50 |
|  |  | 0.03 | 4 | 0.68 | 0.04 | 5.09 | 0.00 | -15 | 3.5 | 68 |
|  |  | 0.03 | 8 | 0.54 | 0.04 | 5.08 | 0.00 | -1.5 | 42.5 | -7 |
|  |  | 0.04 | 1 | 0.86 | 0.05 | 5.05 | 0.00 | -12 | -34 | -1 |
| FORAGING PATCH PHASE |  |  |  |  |  |  |  |  |  |  |
| set |  | cluster |  |  | peak |  |  | MNI coordinates |  |  |
| p | c | p<sub>FWE-corr</sub> | k<sub>E</sub> | p<sub>uncorr</sub> | p<sub>FWE-corr</sub> | T | p<sub>uncorr</sub> | x | y | z |
| 0.00 | 17 | 0.00 | 15668 | 0.00 | 0.00 | 9.79 | 0.00 | 42 | -59.5 | -13 |
|  |  | 0.00 | 15553 | 0.00 | 0.00 | 9.25 | 0.00 | -30 | 17 | -8.5 |
|  |  | 0.00 | 2651 | 0.00 | 0.00 | 8.73 | 0.00 | 42 | 6.5 | 29 |
|  |  | 0.00 | 1891 | 0.00 | 0.00 | 7.59 | 0.00 | -25.5 | -52 | 51.5 |
|  |  | 0.00 | 340 | 0.00 | 0.00 | 6.50 | 0.00 | 1.5 | 6.5 | 29 |
|  |  | 0.00 | 307 | 0.00 | 0.00 | 6.36 | 0.00 | -42 | 3.5 | 30.5 |
|  |  | 0.00 | 141 | 0.03 | 0.00 | 6.20 | 0.00 | -10.5 | 6.5 | -2.5 |
|  |  | 0.00 | 309 | 0.00 | 0.00 | 5.95 | 0.00 | -28.5 | -34 | -38.5 |
|  |  | 0.02 | 23 | 0.34 | 0.01 | 5.41 | 0.00 | 22.5 | -28 | -4 |
|  |  | 0.00 | 153 | 0.02 | 0.01 | 5.40 | 0.00 | -21 | -28 | 20 |
|  |  | 0.02 | 13 | 0.48 | 0.02 | 5.29 | 0.00 | -19.5 | 39.5 | -10 |
|  |  | 0.03 | 7 | 0.61 | 0.03 | 5.14 | 0.00 | 15 | -5.5 | -5.5 |
|  |  | 0.03 | 11 | 0.52 | 0.03 | 5.14 | 0.00 | -54 | 11 | 39.5 |
|  |  | 0.03 | 7 | 0.61 | 0.04 | 5.08 | 0.00 | 13.5 | -26.5 | 21.5 |
|  |  | 0.03 | 6 | 0.64 | 0.04 | 5.06 | 0.00 | -28.5 | -1 | 50 |
|  |  | 0.04 | 3 | 0.76 | 0.04 | 5.06 | 0.00 | 0 | -53.5 | -34 |
|  |  | 0.03 | 6 | 0.64 | 0.04 | 5.04 | 0.00 | -27 | -38.5 | 9.5 |

*Note*. MNI: Montreal Neurological Institute.

### Reward condition x group x time interaction

**Supplementary Table S2.** Reward condition x group x time interaction on *p* < .01.

| **set** |  | **cluster** |  |  | **peak** |  |  | **MNI coordinates** |  |  |
| **p** | **c** | **p<sub>FWE-corr</sub>** | **k<sub>E</sub>** | **p<sub>uncorr</sub>** | **p<sub>FWE-corr</sub>** | **T** | **p<sub>uncorr</sub>** | **x** | **y** | **z** |
|---|---|---|---|---|---|---|---|---|---|---|
| 0.00 | 61 | 0.01 | 3730.00 | 0.00 | 0.10 | 4.81 | 0.00 | 67.5 | -13 | -25 |
|  |  | 0.42 | 1429.00 | 0.02 | 0.53 | 4.15 | 0.00 | -39 | -32.5 | -19 |
|  |  | 1.00 | 238.00 | 0.29 | 0.78 | 3.89 | 0.00 | -24 | -53.5 | -34 |
|  |  | 0.13 | 2212.00 | 0.00 | 0.98 | 3.48 | 0.00 | -16.5 | -5.5 | -35.5 |
|  |  | 1.00 | 167.00 | 0.38 | 0.99 | 3.42 | 0.00 | -24 | 36.5 | -16 |
|  |  | 1.00 | 203.00 | 0.33 | 0.99 | 3.38 | 0.00 | 33 | 60.5 | -16 |
|  |  | 1.00 | 99.00 | 0.50 | 1.00 | 3.36 | 0.00 | -33 | -64 | -32.5 |
|  |  | 1.00 | 200.00 | 0.33 | 1.00 | 3.32 | 0.00 | 24 | 8 | -14.5 |
|  |  | 1.00 | 114.00 | 0.47 | 1.00 | 3.26 | 0.00 | -40.5 | 29 | -19 |
|  |  | 1.00 | 191.00 | 0.34 | 1.00 | 3.25 | 0.00 | 10.5 | 2 | -32.5 |
|  |  | 0.64 | 1113.00 | 0.03 | 1.00 | 3.19 | 0.00 | 7.5 | -95.5 | -2.5 |
|  |  | 0.97 | 576.00 | 0.11 | 1.00 | 3.19 | 0.00 | -18 | -10 | -22 |
|  |  | 0.99 | 454.00 | 0.15 | 1.00 | 3.17 | 0.00 | 34.5 | -37 | -22 |
|  |  | 1.00 | 311.00 | 0.23 | 1.00 | 3.14 | 0.00 | -9 | -76 | -8.5 |
|  |  | 0.97 | 577.00 | 0.11 | 1.00 | 3.14 | 0.00 | 28.5 | -26.5 | -44.5 |
|  |  | 1.00 | 53.00 | 0.64 | 1.00 | 3.11 | 0.00 | -1.5 | -62.5 | 68 |
|  |  | 1.00 | 140.00 | 0.42 | 1.00 | 3.08 | 0.00 | -30 | -46 | -20.5 |
|  |  | 1.00 | 203.00 | 0.33 | 1.00 | 3.05 | 0.00 | 22.5 | -13 | -23.5 |
|  |  | 1.00 | 41.00 | 0.68 | 1.00 | 2.97 | 0.00 | 34.5 | 26 | -20.5 |
|  |  | 1.00 | 278.00 | 0.25 | 1.00 | 2.95 | 0.00 | -37.5 | -68.5 | -11.5 |
|  |  | 1.00 | 125.00 | 0.45 | 1.00 | 2.86 | 0.00 | 39 | -20.5 | -7 |
|  |  | 1.00 | 53.00 | 0.64 | 1.00 | 2.83 | 0.00 | 48 | -68.5 | -10 |
|  |  | 1.00 | 103.00 | 0.49 | 1.00 | 2.83 | 0.00 | -43.5 | -55 | -37 |
|  |  | 1.00 | 42.00 | 0.68 | 1.00 | 2.81 | 0.00 | 34.5 | -20.5 | 8 |
|  |  | 1.00 | 116.00 | 0.47 | 1.00 | 2.78 | 0.00 | -39 | -80.5 | -7 |
|  |  | 1.00 | 25.00 | 0.76 | 1.00 | 2.75 | 0.00 | -9 | -74.5 | 21.5 |
|  |  | 1.00 | 108.00 | 0.48 | 1.00 | 2.74 | 0.00 | 49.5 | -52 | -16 |
|  |  | 1.00 | 48.00 | 0.65 | 1.00 | 2.72 | 0.00 | 27 | -55 | -14.5 |
|  |  | 1.00 | 39.00 | 0.69 | 1.00 | 2.71 | 0.00 | 58.5 | -16 | 47 |
|  |  | 1.00 | 30.00 | 0.73 | 1.00 | 2.67 | 0.00 | 7.5 | -53.5 | -31 |
|  |  | 1.00 | 47.00 | 0.66 | 1.00 | 2.67 | 0.00 | -60 | -4 | 11 |
|  |  | 1.00 | 31.00 | 0.73 | 1.00 | 2.64 | 0.01 | 42 | -32.5 | 27.5 |
|  |  | 1.00 | 12.00 | 0.85 | 1.00 | 2.64 | 0.01 | -27 | -14.5 | 41 |
|  |  | 1.00 | 4.00 | 0.92 | 1.00 | 2.63 | 0.01 | 7.5 | -11.5 | -56.5 |
|  |  | 1.00 | 41.00 | 0.68 | 1.00 | 2.60 | 0.01 | -21 | -37 | -32.5 |
|  |  | 1.00 | 11.00 | 0.85 | 1.00 | 2.60 | 0.01 | 18 | -7 | 35 |
|  |  | 1.00 | 12.00 | 0.85 | 1.00 | 2.60 | 0.01 | 0 | 6.5 | -22 |
|  |  | 1.00 | 11.00 | 0.85 | 1.00 | 2.59 | 0.01 | -1.5 | -91 | 36.5 |
|  |  | 1.00 | 13.00 | 0.84 | 1.00 | 2.58 | 0.01 | -37.5 | -38.5 | 14 |
|  |  | 1.00 | 9.00 | 0.87 | 1.00 | 2.56 | 0.01 | -9 | 72.5 | 12.5 |
|  |  | 1.00 | 14.00 | 0.83 | 1.00 | 2.55 | 0.01 | 64.5 | -8.5 | 18.5 |
|  |  | 1.00 | 21.00 | 0.78 | 1.00 | 2.53 | 0.01 | 42 | -23.5 | 21.5 |
|  |  | 1.00 | 11.00 | 0.85 | 1.00 | 2.52 | 0.01 | 13.5 | -95.5 | 14 |
|  |  | 1.00 | 7.00 | 0.89 | 1.00 | 2.49 | 0.01 | 48 | -44.5 | -2.5 |
|  |  | 1.00 | 13.00 | 0.84 | 1.00 | 2.49 | 0.01 | 19.5 | -58 | -25 |
|  |  | 1.00 | 4.00 | 0.92 | 1.00 | 2.48 | 0.01 | 48 | -49 | -46 |
|  |  | 1.00 | 8.00 | 0.88 | 1.00 | 2.47 | 0.01 | 1.5 | 11 | -16 |
|  |  | 1.00 | 8.00 | 0.88 | 1.00 | 2.47 | 0.01 | -6 | 12.5 | 20 |
|  |  | 1.00 | 2.00 | 0.95 | 1.00 | 2.46 | 0.01 | -63 | -25 | 0.5 |
|  |  | 1.00 | 5.00 | 0.91 | 1.00 | 2.44 | 0.01 | -24 | -17.5 | 27.5 |
|  |  | 1.00 | 2.00 | 0.95 | 1.00 | 2.42 | 0.01 | -31.5 | -8.5 | -11.5 |
|  |  | 1.00 | 3.00 | 0.94 | 1.00 | 2.42 | 0.01 | -3 | 18.5 | -25 |
|  |  | 1.00 | 1.00 | 0.97 | 1.00 | 2.41 | 0.01 | -4.5 | 32 | -34 |
|  |  | 1.00 | 3.00 | 0.94 | 1.00 | 2.41 | 0.01 | -4.5 | 63.5 | -13 |
|  |  | 1.00 | 2.00 | 0.95 | 1.00 | 2.40 | 0.01 | 0 | 29 | -29.5 |
|  |  | 1.00 | 1.00 | 0.97 | 1.00 | 2.39 | 0.01 | 16.5 | 23 | -19 |
|  |  | 1.00 | 1.00 | 0.97 | 1.00 | 2.39 | 0.01 | -67.5 | -44.5 | 2 |
|  |  | 1.00 | 3.00 | 0.94 | 1.00 | 2.39 | 0.01 | 39 | -5.5 | 9.5 |
|  |  | 1.00 | 1.00 | 0.97 | 1.00 | 2.39 | 0.01 | 72 | -25 | 8 |
|  |  | 1.00 | 1.00 | 0.97 | 1.00 | 2.39 | 0.01 | -15 | 24.5 | -22 |
|  |  | 1.00 | 1.00 | 0.97 | 1.00 | 2.39 | 0.01 | -19.5 | -58 | -23.5 |

*Note*. MNI: Montreal Neurological Institute.

### Cue + Foraging Phase

**Supplementary Table S3.** Cue + foraging phase on p < .01 threshold.

| **set** |  | **cluster** |  |  | **peak** |  |  | **MNI coordinates** |  |  |
|---|---|---|---|---|---|---|---|---|---|---|
| **p** | **c** | **p<sub>FWE-corr</sub>** | **k<sub>E</sub>** | **p<sub>uncorr</sub>** | **p<sub>FWE-corr</sub>** | **T** | **p<sub>uncorr</sub>** | **x** | **y** | **z** |
| 0.99 | 229 | 0.14 | 349.00 | 0.00 | 0.65 | 4.78 | 0.00 | 36 | 45.5 | 8 |
|  |  | 0.75 | 205.00 | 0.01 | 0.96 | 4.40 | 0.00 | 63 | -19 | -19 |
|  |  | 1.00 | 54.00 | 0.12 | 1.00 | 4.12 | 0.00 | 39 | -37 | 26 |
|  |  | 0.95 | 160.00 | 0.01 | 1.00 | 4.05 | 0.00 | 61.5 | -40 | -13 |
|  |  | 1.00 | 41.00 | 0.17 | 1.00 | 3.93 | 0.00 | 34.5 | 42.5 | -14.5 |
|  |  | 1.00 | 95.00 | 0.04 | 1.00 | 3.88 | 0.00 | -43.5 | 57.5 | 3.5 |
|  |  | 1.00 | 76.00 | 0.07 | 1.00 | 3.83 | 0.00 | 16.5 | -71.5 | 32 |
|  |  | 1.00 | 35.00 | 0.20 | 1.00 | 3.83 | 0.00 | -24 | -59.5 | 29 |
|  |  | 1.00 | 35.00 | 0.20 | 1.00 | 3.82 | 0.00 | -25.5 | -64 | 6.5 |
|  |  | 1.00 | 22.00 | 0.31 | 1.00 | 3.78 | 0.00 | 0 | 21.5 | 5 |
|  |  | 1.00 | 40.00 | 0.17 | 1.00 | 3.70 | 0.00 | -28.5 | 39.5 | -13 |
|  |  | 0.67 | 217.00 | 0.00 | 1.00 | 3.69 | 0.00 | 34.5 | 59 | -11.5 |
|  |  | 1.00 | 39.00 | 0.18 | 1.00 | 3.64 | 0.00 | 45 | -23.5 | -19 |
|  |  | 1.00 | 61.00 | 0.10 | 1.00 | 3.60 | 0.00 | 18 | -53.5 | 27.5 |
|  |  | 1.00 | 18.00 | 0.35 | 1.00 | 3.59 | 0.00 | -31.5 | -49 | 30.5 |
|  |  | 1.00 | 7.00 | 0.58 | 1.00 | 3.52 | 0.00 | -66 | -11.5 | -20.5 |
|  |  | 1.00 | 29.00 | 0.24 | 1.00 | 3.52 | 0.00 | 7.5 | -28 | 30.5 |
|  |  | 1.00 | 25.00 | 0.27 | 1.00 | 3.47 | 0.00 | 24 | 6.5 | 21.5 |
|  |  | 1.00 | 33.00 | 0.21 | 1.00 | 3.46 | 0.00 | 42 | 6.5 | -11.5 |
|  |  | 1.00 | 56.00 | 0.11 | 1.00 | 3.45 | 0.00 | -1.5 | -31 | -8.5 |
|  |  | 1.00 | 40.00 | 0.17 | 1.00 | 3.35 | 0.00 | -13.5 | -62.5 | 33.5 |
|  |  | 1.00 | 18.00 | 0.35 | 1.00 | 3.31 | 0.00 | -36 | 62 | 8 |
|  |  | 1.00 | 50.00 | 0.13 | 1.00 | 3.25 | 0.00 | -12 | -11.5 | -29.5 |
|  |  | 1.00 | 90.00 | 0.05 | 1.00 | 3.24 | 0.00 | 7.5 | -97 | -5.5 |
|  |  | 1.00 | 61.00 | 0.10 | 1.00 | 3.23 | 0.00 | -15 | -91 | 15.5 |
|  |  | 1.00 | 31.00 | 0.23 | 1.00 | 3.21 | 0.00 | -13.5 | -86.5 | 9.5 |
|  |  | 1.00 | 69.00 | 0.08 | 1.00 | 3.21 | 0.00 | 46.5 | -71.5 | 42.5 |
|  |  | 1.00 | 11.00 | 0.47 | 1.00 | 3.21 | 0.00 | 45 | -17.5 | -28 |
|  |  | 1.00 | 43.00 | 0.16 | 1.00 | 3.20 | 0.00 | 21 | -64 | 23 |
|  |  | 1.00 | 21.00 | 0.32 | 1.00 | 3.18 | 0.00 | -57 | -19 | 3.5 |
|  |  | 1.00 | 20.00 | 0.33 | 1.00 | 3.18 | 0.00 | 46.5 | -52 | -10 |
|  |  | 1.00 | 9.00 | 0.52 | 1.00 | 3.15 | 0.00 | -22.5 | 11 | 32 |
|  |  | 1.00 | 27.00 | 0.26 | 1.00 | 3.14 | 0.00 | -3 | -11.5 | -19 |
|  |  | 1.00 | 84.00 | 0.06 | 1.00 | 3.13 | 0.00 | -63 | -38.5 | -22 |
|  |  | 1.00 | 18.00 | 0.35 | 1.00 | 3.12 | 0.00 | -39 | -29.5 | -19 |
|  |  | 1.00 | 11.00 | 0.47 | 1.00 | 3.10 | 0.00 | 46.5 | -29.5 | 9.5 |
|  |  | 1.00 | 7.00 | 0.58 | 1.00 | 3.09 | 0.00 | 31.5 | 12.5 | 21.5 |
|  |  | 1.00 | 7.00 | 0.58 | 1.00 | 3.09 | 0.00 | 34.5 | 66.5 | 6.5 |
|  |  | 1.00 | 8.00 | 0.55 | 1.00 | 3.07 | 0.00 | 36 | -56.5 | 18.5 |
|  |  | 1.00 | 23.00 | 0.29 | 1.00 | 3.07 | 0.00 | 13.5 | -94 | 14 |
|  |  | 1.00 | 9.00 | 0.52 | 1.00 | 3.06 | 0.00 | -60 | 2 | -22 |
|  |  | 1.00 | 9.00 | 0.52 | 1.00 | 3.03 | 0.00 | 33 | -47.5 | 14 |
|  |  | 1.00 | 33.00 | 0.21 | 1.00 | 3.03 | 0.00 | -33 | -74.5 | 24.5 |
|  |  | 1.00 | 8.00 | 0.55 | 1.00 | 3.03 | 0.00 | 18 | -38.5 | -19 |
|  |  | 1.00 | 12.00 | 0.45 | 1.00 | 3.03 | 0.00 | -36 | -85 | -10 |
|  |  | 1.00 | 12.00 | 0.45 | 1.00 | 3.02 | 0.00 | 4.5 | -41.5 | 21.5 |
|  |  | 1.00 | 14.00 | 0.42 | 1.00 | 3.00 | 0.00 | -25.5 | -53.5 | -34 |
|  |  | 1.00 | 6.00 | 0.61 | 1.00 | 3.00 | 0.00 | 28.5 | -37 | -37 |
|  |  | 1.00 | 13.00 | 0.43 | 1.00 | 3.00 | 0.00 | -43.5 | -22 | -2.5 |
|  |  | 1.00 | 22.00 | 0.31 | 1.00 | 2.97 | 0.00 | -10.5 | -29.5 | -14.5 |
|  |  | 1.00 | 24.00 | 0.28 | 1.00 | 2.96 | 0.00 | 21 | -16 | -25 |
|  |  | 1.00 | 9.00 | 0.52 | 1.00 | 2.96 | 0.00 | 7.5 | -4 | -2.5 |
|  |  | 1.00 | 14.00 | 0.42 | 1.00 | 2.96 | 0.00 | -24 | -25 | -22 |
|  |  | 1.00 | 8.00 | 0.55 | 1.00 | 2.96 | 0.00 | 57 | 6.5 | -34 |
|  |  | 1.00 | 12.00 | 0.45 | 1.00 | 2.96 | 0.00 | 16.5 | 44 | 2 |
|  |  | 1.00 | 6.00 | 0.61 | 1.00 | 2.95 | 0.00 | 46.5 | -2.5 | 23 |
|  |  | 1.00 | 54.00 | 0.12 | 1.00 | 2.95 | 0.00 | -6 | -40 | 18.5 |
|  |  | 1.00 | 10.00 | 0.50 | 1.00 | 2.94 | 0.00 | 25.5 | 11 | -43 |
|  |  | 1.00 | 19.00 | 0.34 | 1.00 | 2.94 | 0.00 | -4.5 | -88 | 11 |
|  |  | 1.00 | 27.00 | 0.26 | 1.00 | 2.93 | 0.00 | 42 | -43 | -2.5 |
|  |  | 1.00 | 32.00 | 0.22 | 1.00 | 2.93 | 0.00 | -27 | -16 | 29 |
|  |  | 1.00 | 33.00 | 0.21 | 1.00 | 2.93 | 0.00 | 42 | -43 | 65 |
|  |  | 1.00 | 17.00 | 0.37 | 1.00 | 2.91 | 0.00 | -55.5 | -7 | -29.5 |
|  |  | 1.00 | 8.00 | 0.55 | 1.00 | 2.90 | 0.00 | 0 | -88 | -17.5 |
|  |  | 1.00 | 21.00 | 0.32 | 1.00 | 2.90 | 0.00 | 28.5 | -56.5 | -14.5 |
|  |  | 1.00 | 9.00 | 0.52 | 1.00 | 2.90 | 0.00 | -51 | -34 | -16 |
|  |  | 1.00 | 9.00 | 0.52 | 1.00 | 2.89 | 0.00 | 31.5 | 21.5 | -7 |
|  |  | 1.00 | 10.00 | 0.50 | 1.00 | 2.89 | 0.00 | 43.5 | -25 | 18.5 |
|  |  | 1.00 | 6.00 | 0.61 | 1.00 | 2.88 | 0.00 | 10.5 | 18.5 | -20.5 |
|  |  | 1.00 | 17.00 | 0.37 | 1.00 | 2.88 | 0.00 | -31.5 | -64 | 62 |
|  |  | 1.00 | 12.00 | 0.45 | 1.00 | 2.87 | 0.00 | -27 | -35.5 | 44 |
|  |  | 1.00 | 2.00 | 0.79 | 1.00 | 2.86 | 0.00 | 63 | 20 | 20 |
|  |  | 1.00 | 8.00 | 0.55 | 1.00 | 2.86 | 0.00 | -4.5 | 24.5 | 2 |
|  |  | 1.00 | 6.00 | 0.61 | 1.00 | 2.85 | 0.00 | 42 | -16 | -17.5 |
|  |  | 1.00 | 1.00 | 0.86 | 1.00 | 2.84 | 0.00 | -64.5 | -52 | 30.5 |
|  |  | 1.00 | 5.00 | 0.64 | 1.00 | 2.82 | 0.00 | 13.5 | -38.5 | 20 |
|  |  | 1.00 | 8.00 | 0.55 | 1.00 | 2.82 | 0.00 | 7.5 | 60.5 | 2 |
|  |  | 1.00 | 5.00 | 0.64 | 1.00 | 2.82 | 0.00 | 36 | 14 | 2 |
|  |  | 1.00 | 2.00 | 0.79 | 1.00 | 2.81 | 0.00 | 10.5 | 9.5 | 59 |
|  |  | 1.00 | 6.00 | 0.61 | 1.00 | 2.80 | 0.00 | -39 | -34 | 8 |
|  |  | 1.00 | 7.00 | 0.58 | 1.00 | 2.80 | 0.00 | -54 | -5.5 | -40 |
|  |  | 1.00 | 4.00 | 0.68 | 1.00 | 2.80 | 0.00 | 66 | -5.5 | -20.5 |
|  |  | 1.00 | 10.00 | 0.50 | 1.00 | 2.80 | 0.00 | -1.5 | -77.5 | -20.5 |
|  |  | 1.00 | 8.00 | 0.55 | 1.00 | 2.80 | 0.00 | 30 | -35.5 | 62 |
|  |  | 1.00 | 17.00 | 0.37 | 1.00 | 2.79 | 0.00 | -34.5 | 62 | -8.5 |
|  |  | 1.00 | 8.00 | 0.55 | 1.00 | 2.79 | 0.00 | 13.5 | -56.5 | 15.5 |
|  |  | 1.00 | 25.00 | 0.27 | 1.00 | 2.79 | 0.00 | -33 | -85 | 14 |
|  |  | 1.00 | 12.00 | 0.45 | 1.00 | 2.79 | 0.00 | 24 | -19 | 20 |
|  |  | 1.00 | 7.00 | 0.58 | 1.00 | 2.78 | 0.00 | 31.5 | 3.5 | -7 |
|  |  | 1.00 | 9.00 | 0.52 | 1.00 | 2.78 | 0.00 | 6 | -56.5 | -14.5 |
|  |  | 1.00 | 9.00 | 0.52 | 1.00 | 2.78 | 0.00 | -19.5 | -73 | 39.5 |
|  |  | 1.00 | 7.00 | 0.58 | 1.00 | 2.77 | 0.00 | -21 | -17.5 | 39.5 |
|  |  | 1.00 | 19.00 | 0.34 | 1.00 | 2.77 | 0.00 | -10.5 | 17 | -17.5 |
|  |  | 1.00 | 9.00 | 0.52 | 1.00 | 2.77 | 0.00 | -40.5 | 50 | -16 |
|  |  | 1.00 | 5.00 | 0.64 | 1.00 | 2.77 | 0.00 | -37.5 | -49 | -16 |
|  |  | 1.00 | 7.00 | 0.58 | 1.00 | 2.77 | 0.00 | 16.5 | 24.5 | 27.5 |
|  |  | 1.00 | 11.00 | 0.47 | 1.00 | 2.76 | 0.00 | 21 | 56 | -17.5 |
|  |  | 1.00 | 33.00 | 0.21 | 1.00 | 2.76 | 0.00 | -15 | 62 | -7 |
|  |  | 1.00 | 7.00 | 0.58 | 1.00 | 2.76 | 0.00 | 22.5 | -91 | 14 |
|  |  | 1.00 | 3.00 | 0.73 | 1.00 | 2.75 | 0.00 | 48 | -35.5 | 38 |
|  |  | 1.00 | 6.00 | 0.61 | 1.00 | 2.75 | 0.00 | -10.5 | -62.5 | 20 |
|  |  | 1.00 | 12.00 | 0.45 | 1.00 | 2.74 | 0.00 | 61.5 | -28 | 35 |
|  |  | 1.00 | 6.00 | 0.61 | 1.00 | 2.74 | 0.00 | 15 | -79 | 20 |
|  |  | 1.00 | 7.00 | 0.58 | 1.00 | 2.74 | 0.00 | -24 | -55 | 17 |
|  |  | 1.00 | 15.00 | 0.40 | 1.00 | 2.73 | 0.00 | -13.5 | -37 | 30.5 |
|  |  | 1.00 | 3.00 | 0.73 | 1.00 | 2.73 | 0.00 | -13.5 | -77.5 | 24.5 |
|  |  | 1.00 | 8.00 | 0.55 | 1.00 | 2.73 | 0.00 | 36 | -7 | -17.5 |
|  |  | 1.00 | 2.00 | 0.79 | 1.00 | 2.73 | 0.00 | -6 | -68.5 | 65 |
|  |  | 1.00 | 4.00 | 0.68 | 1.00 | 2.72 | 0.00 | -52.5 | -49 | -28 |
|  |  | 1.00 | 4.00 | 0.68 | 1.00 | 2.72 | 0.00 | -52.5 | -20.5 | -28 |
|  |  | 1.00 | 6.00 | 0.61 | 1.00 | 2.72 | 0.00 | 1.5 | -71.5 | 47 |
|  |  | 1.00 | 2.00 | 0.79 | 1.00 | 2.71 | 0.00 | 6 | -91 | 18.5 |
|  |  | 1.00 | 8.00 | 0.55 | 1.00 | 2.71 | 0.00 | 51 | -68.5 | -8.5 |
|  |  | 1.00 | 10.00 | 0.50 | 1.00 | 2.70 | 0.00 | -42 | -40 | -22 |
|  |  | 1.00 | 11.00 | 0.47 | 1.00 | 2.70 | 0.00 | 45 | -58 | 32 |
|  |  | 1.00 | 4.00 | 0.68 | 1.00 | 2.70 | 0.00 | 34.5 | 23 | 30.5 |
|  |  | 1.00 | 1.00 | 0.86 | 1.00 | 2.70 | 0.00 | -67.5 | -20.5 | -22 |
|  |  | 1.00 | 8.00 | 0.55 | 1.00 | 2.69 | 0.00 | 10.5 | 24.5 | 11 |
|  |  | 1.00 | 20.00 | 0.33 | 1.00 | 2.68 | 0.00 | 3 | -4 | -29.5 |
|  |  | 1.00 | 3.00 | 0.73 | 1.00 | 2.68 | 0.00 | 16.5 | 3.5 | -35.5 |
|  |  | 1.00 | 4.00 | 0.68 | 1.00 | 2.68 | 0.00 | -27 | -37 | -35.5 |
|  |  | 1.00 | 1.00 | 0.86 | 1.00 | 2.68 | 0.00 | 28.5 | -7 | 6.5 |
|  |  | 1.00 | 11.00 | 0.47 | 1.00 | 2.68 | 0.00 | -36 | 9.5 | 24.5 |
|  |  | 1.00 | 5.00 | 0.64 | 1.00 | 2.68 | 0.00 | 12 | 29 | 45.5 |
|  |  | 1.00 | 6.00 | 0.61 | 1.00 | 2.67 | 0.00 | -27 | -10 | -1 |
|  |  | 1.00 | 2.00 | 0.79 | 1.00 | 2.67 | 0.00 | 39 | -20.5 | 30.5 |
|  |  | 1.00 | 2.00 | 0.79 | 1.00 | 2.67 | 0.00 | 7.5 | 44 | -23.5 |
|  |  | 1.00 | 13.00 | 0.43 | 1.00 | 2.66 | 0.00 | 39 | -35.5 | -17.5 |
|  |  | 1.00 | 2.00 | 0.79 | 1.00 | 2.66 | 0.00 | -31.5 | -89.5 | -7 |
|  |  | 1.00 | 2.00 | 0.79 | 1.00 | 2.65 | 0.01 | -49.5 | -55 | -43 |
|  |  | 1.00 | 6.00 | 0.61 | 1.00 | 2.65 | 0.01 | -18 | 15.5 | 41 |
|  |  | 1.00 | 1.00 | 0.86 | 1.00 | 2.65 | 0.01 | 51 | 50 | 5 |
|  |  | 1.00 | 4.00 | 0.68 | 1.00 | 2.65 | 0.01 | 4.5 | -7 | 51.5 |
|  |  | 1.00 | 7.00 | 0.58 | 1.00 | 2.64 | 0.01 | -12 | -97 | 14 |
|  |  | 1.00 | 5.00 | 0.64 | 1.00 | 2.64 | 0.01 | -12 | 8 | 47 |
|  |  | 1.00 | 4.00 | 0.68 | 1.00 | 2.64 | 0.01 | 19.5 | 47 | -17.5 |
|  |  | 1.00 | 12.00 | 0.45 | 1.00 | 2.64 | 0.01 | 31.5 | -68.5 | 53 |
|  |  | 1.00 | 7.00 | 0.58 | 1.00 | 2.63 | 0.01 | 19.5 | -47.5 | -34 |
|  |  | 1.00 | 5.00 | 0.64 | 1.00 | 2.62 | 0.01 | -31.5 | -49 | 18.5 |
|  |  | 1.00 | 5.00 | 0.64 | 1.00 | 2.62 | 0.01 | -10.5 | 24.5 | -17.5 |
|  |  | 1.00 | 2.00 | 0.79 | 1.00 | 2.62 | 0.01 | 36 | 11 | 20 |
|  |  | 1.00 | 3.00 | 0.73 | 1.00 | 2.61 | 0.01 | -24 | -4 | 14 |
|  |  | 1.00 | 7.00 | 0.58 | 1.00 | 2.61 | 0.01 | 22.5 | 35 | -17.5 |
|  |  | 1.00 | 3.00 | 0.73 | 1.00 | 2.60 | 0.01 | -19.5 | 41 | -16 |
|  |  | 1.00 | 3.00 | 0.73 | 1.00 | 2.60 | 0.01 | 27 | 41 | -8.5 |
|  |  | 1.00 | 7.00 | 0.58 | 1.00 | 2.60 | 0.01 | 10.5 | -74.5 | 44 |
|  |  | 1.00 | 9.00 | 0.52 | 1.00 | 2.60 | 0.01 | 33 | -59.5 | -19 |
|  |  | 1.00 | 6.00 | 0.61 | 1.00 | 2.59 | 0.01 | -27 | -76 | 42.5 |
|  |  | 1.00 | 3.00 | 0.73 | 1.00 | 2.59 | 0.01 | 27 | 17 | -25 |
|  |  | 1.00 | 2.00 | 0.79 | 1.00 | 2.59 | 0.01 | 51 | -23.5 | 59 |
|  |  | 1.00 | 2.00 | 0.79 | 1.00 | 2.59 | 0.01 | -15 | -16 | 3.5 |
|  |  | 1.00 | 5.00 | 0.64 | 1.00 | 2.59 | 0.01 | -13.5 | -25 | 15.5 |
|  |  | 1.00 | 2.00 | 0.79 | 1.00 | 2.59 | 0.01 | -36 | -56.5 | -38.5 |
|  |  | 1.00 | 1.00 | 0.86 | 1.00 | 2.57 | 0.01 | 28.5 | -50.5 | 35 |
|  |  | 1.00 | 3.00 | 0.73 | 1.00 | 2.57 | 0.01 | 58.5 | -61 | -23.5 |
|  |  | 1.00 | 6.00 | 0.61 | 1.00 | 2.57 | 0.01 | 0 | 3.5 | -19 |
|  |  | 1.00 | 2.00 | 0.79 | 1.00 | 2.57 | 0.01 | 31.5 | -91 | 6.5 |
|  |  | 1.00 | 7.00 | 0.58 | 1.00 | 2.57 | 0.01 | 16.5 | 29 | -19 |
|  |  | 1.00 | 1.00 | 0.86 | 1.00 | 2.57 | 0.01 | 64.5 | -23.5 | -7 |
|  |  | 1.00 | 4.00 | 0.68 | 1.00 | 2.56 | 0.01 | 1.5 | -43 | -28 |
|  |  | 1.00 | 2.00 | 0.79 | 1.00 | 2.56 | 0.01 | 6 | -76 | 12.5 |
|  |  | 1.00 | 2.00 | 0.79 | 1.00 | 2.55 | 0.01 | -4.5 | -13 | 2 |
|  |  | 1.00 | 4.00 | 0.68 | 1.00 | 2.55 | 0.01 | -37.5 | -61 | 44 |
|  |  | 1.00 | 6.00 | 0.61 | 1.00 | 2.54 | 0.01 | -3 | -59.5 | -10 |
|  |  | 1.00 | 3.00 | 0.73 | 1.00 | 2.54 | 0.01 | 7.5 | -70 | 35 |
|  |  | 1.00 | 3.00 | 0.73 | 1.00 | 2.54 | 0.01 | -49.5 | -29.5 | 5 |
|  |  | 1.00 | 1.00 | 0.86 | 1.00 | 2.54 | 0.01 | 31.5 | -40 | -22 |
|  |  | 1.00 | 2.00 | 0.79 | 1.00 | 2.54 | 0.01 | -30 | -5.5 | 18.5 |
|  |  | 1.00 | 4.00 | 0.68 | 1.00 | 2.53 | 0.01 | -31.5 | -44.5 | -44.5 |
|  |  | 1.00 | 3.00 | 0.73 | 1.00 | 2.53 | 0.01 | 0 | -35.5 | -25 |
|  |  | 1.00 | 1.00 | 0.86 | 1.00 | 2.53 | 0.01 | 28.5 | 24.5 | 17 |
|  |  | 1.00 | 8.00 | 0.55 | 1.00 | 2.53 | 0.01 | 51 | 9.5 | 20 |
|  |  | 1.00 | 8.00 | 0.55 | 1.00 | 2.53 | 0.01 | 33 | -46 | 23 |
|  |  | 1.00 | 2.00 | 0.79 | 1.00 | 2.53 | 0.01 | 7.5 | -83.5 | 3.5 |
|  |  | 1.00 | 1.00 | 0.86 | 1.00 | 2.52 | 0.01 | 25.5 | -1 | 3.5 |
|  |  | 1.00 | 1.00 | 0.86 | 1.00 | 2.52 | 0.01 | 34.5 | 41 | 2 |
|  |  | 1.00 | 1.00 | 0.86 | 1.00 | 2.52 | 0.01 | 1.5 | 56 | -31 |
|  |  | 1.00 | 3.00 | 0.73 | 1.00 | 2.51 | 0.01 | -3 | -83.5 | 14 |
|  |  | 1.00 | 2.00 | 0.79 | 1.00 | 2.51 | 0.01 | -25.5 | -8.5 | -49 |
|  |  | 1.00 | 1.00 | 0.86 | 1.00 | 2.51 | 0.01 | 63 | -59.5 | 30.5 |
|  |  | 1.00 | 2.00 | 0.79 | 1.00 | 2.51 | 0.01 | 16.5 | 9.5 | -28 |
|  |  | 1.00 | 5.00 | 0.64 | 1.00 | 2.50 | 0.01 | -34.5 | -56.5 | 35 |
|  |  | 1.00 | 3.00 | 0.73 | 1.00 | 2.50 | 0.01 | 27 | -55 | -20.5 |
|  |  | 1.00 | 2.00 | 0.79 | 1.00 | 2.50 | 0.01 | -28.5 | 51.5 | -14.5 |
|  |  | 1.00 | 1.00 | 0.86 | 1.00 | 2.50 | 0.01 | -16.5 | -11.5 | -7 |
|  |  | 1.00 | 5.00 | 0.64 | 1.00 | 2.49 | 0.01 | 58.5 | -17.5 | 12.5 |
|  |  | 1.00 | 1.00 | 0.86 | 1.00 | 2.49 | 0.01 | 40.5 | -40 | 5 |
|  |  | 1.00 | 2.00 | 0.79 | 1.00 | 2.49 | 0.01 | 33 | -34 | 6.5 |
|  |  | 1.00 | 2.00 | 0.79 | 1.00 | 2.48 | 0.01 | 45 | -4 | 8 |
|  |  | 1.00 | 1.00 | 0.86 | 1.00 | 2.48 | 0.01 | 52.5 | -56.5 | -14.5 |
|  |  | 1.00 | 1.00 | 0.86 | 1.00 | 2.48 | 0.01 | -21 | -82 | -32.5 |
|  |  | 1.00 | 2.00 | 0.79 | 1.00 | 2.47 | 0.01 | 37.5 | 17 | -16 |
|  |  | 1.00 | 5.00 | 0.64 | 1.00 | 2.47 | 0.01 | -9 | -94 | -4 |
|  |  | 1.00 | 2.00 | 0.79 | 1.00 | 2.47 | 0.01 | -24 | -86.5 | 0.5 |
|  |  | 1.00 | 1.00 | 0.86 | 1.00 | 2.46 | 0.01 | 16.5 | 72.5 | -1 |
|  |  | 1.00 | 3.00 | 0.73 | 1.00 | 2.46 | 0.01 | 12 | 62 | -4 |
|  |  | 1.00 | 1.00 | 0.86 | 1.00 | 2.46 | 0.01 | -40.5 | -79 | -13 |
|  |  | 1.00 | 2.00 | 0.79 | 1.00 | 2.46 | 0.01 | 69 | -2.5 | 12.5 |
|  |  | 1.00 | 1.00 | 0.86 | 1.00 | 2.46 | 0.01 | 34.5 | 0.5 | -20.5 |
|  |  | 1.00 | 1.00 | 0.86 | 1.00 | 2.46 | 0.01 | 34.5 | -40 | 17 |
|  |  | 1.00 | 2.00 | 0.79 | 1.00 | 2.45 | 0.01 | -33 | -40 | 24.5 |
|  |  | 1.00 | 1.00 | 0.86 | 1.00 | 2.45 | 0.01 | -25.5 | -10 | 38 |
|  |  | 1.00 | 1.00 | 0.86 | 1.00 | 2.45 | 0.01 | -13.5 | -4 | -49 |
|  |  | 1.00 | 1.00 | 0.86 | 1.00 | 2.44 | 0.01 | 48 | -13 | -31 |
|  |  | 1.00 | 2.00 | 0.79 | 1.00 | 2.44 | 0.01 | 51 | -41.5 | -4 |
|  |  | 1.00 | 1.00 | 0.86 | 1.00 | 2.44 | 0.01 | 21 | -80.5 | 54.5 |
|  |  | 1.00 | 2.00 | 0.79 | 1.00 | 2.43 | 0.01 | 9 | 27.5 | 0.5 |
|  |  | 1.00 | 2.00 | 0.79 | 1.00 | 2.43 | 0.01 | 31.5 | -29.5 | -31 |
|  |  | 1.00 | 1.00 | 0.86 | 1.00 | 2.43 | 0.01 | 9 | -58 | 32 |
|  |  | 1.00 | 1.00 | 0.86 | 1.00 | 2.43 | 0.01 | 18 | -80.5 | 23 |
|  |  | 1.00 | 2.00 | 0.79 | 1.00 | 2.42 | 0.01 | -36 | -49 | -22 |
|  |  | 1.00 | 1.00 | 0.86 | 1.00 | 2.42 | 0.01 | -10.5 | 47 | -19 |
|  |  | 1.00 | 1.00 | 0.86 | 1.00 | 2.42 | 0.01 | 13.5 | 24.5 | 47 |
|  |  | 1.00 | 4.00 | 0.68 | 1.00 | 2.41 | 0.01 | -45 | -46 | -37 |
|  |  | 1.00 | 1.00 | 0.86 | 1.00 | 2.41 | 0.01 | 3 | 20 | -14.5 |
|  |  | 1.00 | 1.00 | 0.86 | 1.00 | 2.41 | 0.01 | 24 | 2 | -26.5 |
|  |  | 1.00 | 1.00 | 0.86 | 1.00 | 2.41 | 0.01 | 33 | 2 | -16 |
|  |  | 1.00 | 1.00 | 0.86 | 1.00 | 2.41 | 0.01 | 10.5 | -14.5 | 30.5 |
|  |  | 1.00 | 1.00 | 0.86 | 1.00 | 2.41 | 0.01 | 3 | -94 | 8 |
|  |  | 1.00 | 1.00 | 0.86 | 1.00 | 2.40 | 0.01 | 72 | -23.5 | 11 |
|  |  | 1.00 | 1.00 | 0.86 | 1.00 | 2.40 | 0.01 | 43.5 | 33.5 | 38 |
|  |  | 1.00 | 1.00 | 0.86 | 1.00 | 2.40 | 0.01 | -48 | -47.5 | -19 |
|  |  | 1.00 | 1.00 | 0.86 | 1.00 | 2.40 | 0.01 | 1.5 | 27.5 | -25 |
|  |  | 1.00 | 1.00 | 0.86 | 1.00 | 2.40 | 0.01 | 39 | -25 | -8.5 |
|  |  | 1.00 | 1.00 | 0.86 | 1.00 | 2.39 | 0.01 | -37.5 | 24.5 | 38 |
|  |  | 1.00 | 1.00 | 0.86 | 1.00 | 2.39 | 0.01 | -40.5 | -23.5 | -10 |
|  |  | 1.00 | 1.00 | 0.86 | 1.00 | 2.39 | 0.01 | 1.5 | -16 | 80 |
|  |  | 1.00 | 1.00 | 0.86 | 1.00 | 2.39 | 0.01 | 54 | -10 | -25 |
|  |  | 1.00 | 1.00 | 0.86 | 1.00 | 2.39 | 0.01 | 7.5 | -70 | -26.5 |

*Note*. MNI: Montreal Neurological Institute.

## References

1. Feighner, J. P. et al. Diagnostic criteria for use in psychiatric research. Arch. Gen. Psychiatry 26, 57–63 (1972).

2. Pizzagalli, D. A. Toward a better understanding of the mechanisms and pathophysiology of anhedonia: Are we ready for translation? Am. J. Psychiatry 179, 458–469 (2022).

3. Abler, B., Grön, G., Hartmann, A., Metzger, C. & Walter, M. Modulation of frontostriatal interaction aligns with reduced primary reward processing under serotonergic drugs. J. Neurosci. 32, 1329–1335 (2012).

4. Daw, N. D., Kakade, S. & Dayan, P. Opponent interactions between serotonin and dopamine. Neural Netw. 15, 603–616 (2002).

5. Soubrié, P. Reconciling the role of central serotonin neurons in human and animal behavior. Behav. Brain Sci. 9, 319–335 (1986).

6. Kranz, G. S., Kasper, S. & Lanzenberger, R. Reward and the serotonergic system. Neuroscience 166, 1023–1035 (2010).

7. Calizo, L. H. et al. Raphe serotonin neurons are not homogenous: Electrophysiological, morphological and neurochemical evidence. Neuropharmacology 61, 524–543 (2011).

8. Ren, J. et al. Single-cell transcriptomes and whole-brain projections of serotonin neurons in the mouse dorsal and median raphe nuclei. eLife 8, e49424 (2019).

9. Serretti, A. Anhedonia: Current and future treatments. Psychiatry Clin. Neurosci. Rep. 4, e70088 (2025).

10. Wu, C., Mu, Q., Gao, W. & Lu, S. The characteristics of anhedonia in depression: a review from a clinically oriented perspective. Transl. Psychiatry 15, 90 (2025).

11. Treadway, M. T. & Zald, D. H. Reconsidering anhedonia in depression: Lessons from translational neuroscience. Neurosci. Biobehav. Rev. 35, 537–555 (2011).

12. Koolschijn, R. S. et al. Resources, costs and long-term value: an integrative perspective on serotonin and meta-decision making. Curr. Opin. Behav. Sci. 60, 101453 (2024).

13. Insel, T. et al. Research Domain Criteria (RDoC): Toward a new classification framework for research on mental disorders. Am. J. Psychiatry 167, 748–751 (2010).

14. Salamone, J. D. & Correa, M. The mysterious motivational functions of mesolimbic dopamine. Neuron 76, 470–485 (2012).

15. Treadway, M. T., Buckholtz, J. W., Schwartzman, A. N., Lambert, W. E. & Zald, D. H. Worth the ‘EEfRT’? The effort expenditure for rewards task as an objective measure of motivation and anhedonia. PLoS ONE 4, e6598 (2009).

16. Treadway, M. T., Bossaller, N. A., Shelton, R. C. & Zald, D. H. Effort-based decision-making in major depressive disorder: A translational model of motivational anhedonia. J. Abnorm. Psychol. 121, 553–558 (2012).

17. Ang, Y.-S., Gelda, S. E. & Pizzagalli, D. A. Cognitive effort-based decision-making in major depressive disorder. Psychol. Med. 53, 4228–4235 (2023).

18. Bustamante, L. A. et al. Major depression symptom severity associations with willingness to exert effort and patch foraging strategy. Preprint at 10.1101/2024.02.18.24302985 (2024).

19. Dong, X., Cooper, J. A., Moran, E. K. & Barch, D. M. Understanding effort-cost decision-making mechanisms in mood and psychotic disorders: a computational modeling approach across physical and cognitive effort paradigms. Biol. Psychiatry Cogn. Neurosci. Neuroimaging 10.1016/j.bpsc.2025.11.004 (2025).

20. Sahni, A., Frey, A.-L. & McCabe, C. Anhedonia is associated with computational impairments in reward and effort learning in young people with depression symptoms. Psychol. Med. 55, e347 (2025).

21. Vinckier, F. et al. Elevated effort cost identified by computational modeling as a distinctive feature explaining multiple behaviors in patients with depression. Biol. Psychiatry Cogn. Neurosci. Neuroimaging 7, 1158–1169 (2022).

22. Burke, W. J. Escitalopram. Expert Opin. Investig. Drugs 11, 1477–1486 (2002).

23. Orsini, C., Bosch, J. E., Labek, K. & Viviani, R. Functional imaging of time on task and the involvement of dopaminergic and cholinergic substrates in cognitive effort and reward. Sci. Rep. 16, 7898 (2026).

24. Viviani, R. et al. Signals of anticipation of reward and of mean reward rates in the human brain. Sci. Rep. 10, 4287 (2020).

25. Schultz, W., Dayan, P. & Montague, P. R. A neural substrate of prediction and reward. Science 275, 1593–1599 (1997).

26. Abler, B., Walter, H., Erk, S., Kammerer, H. & Spitzer, M. Prediction error as a linear function of reward probability is coded in human nucleus accumbens. NeuroImage 31, 790–795 (2006).

27. Walton, M. E. & Bouret, S. What is the relationship between dopamine and effort? Trends Neurosci. 42, 79–91 (2019).

28. Harkin, E. F., Grossman, C. D., Cohen, J. Y., Béïque, J.-C. & Naud, R. A prospective code for value in the serotonin system. Nature 641, 952–959 (2025).

29. Lottem, E. et al. Activation of serotonin neurons promotes active persistence in a probabilistic foraging task. Nat. Commun. 9, 1000 (2018).

30. Berridge, K. C. & O’Doherty, J. P. From experienced utility to decision utility. in Neuroeconomics 2nd edn (eds Glimcher, P. W. & Fehr, E.) 335–351 (Academic Press, 2014).

31. McClure, S. M., Daw, N. D. & Montague, P. R. A computational substrate for incentive salience. Trends Neurosci. 26, 423–428 (2003).

32. Kurniawan, I. T. et al. Choosing to make an effort: The role of striatum in signaling physical effort of a chosen action. J. Neurophysiol. 104, 313–321 (2010).

33. Orsini, C., Huber, D. A., Labek, K., Bosch, J. E. & Viviani, R. Basal forebrain and neural correlates of self-regulation traits in sustained attention. Imaging Neurosci. 4, IMAG.a.1187 (2026).

34. Hamid, A. A. et al. Mesolimbic dopamine signals the value of work. Nat. Neurosci. 19, 117–126 (2016).

35. Morris, L. S. et al. Targeting the dopaminergic midbrain with precision 7-Tesla biofeedback training in depression: a proof-of-principle randomized controlled trial. Mol. Psychiatry 10.1038/s41380-026-03911-x (2026).

36. Husain, M. & Roiser, J. P. Neuroscience of apathy and anhedonia: a transdiagnostic approach. Nat. Rev. Neurosci. 19, 470–484 (2018).

37. Berridge, K. C., Robinson, T. E. & Aldridge, J. W. Dissecting components of reward: ‘liking’, ‘wanting’, and learning. Curr. Opin. Pharmacol. 9, 65–73 (2009).

38. Krystal, A. D. et al. A randomized proof-of-mechanism trial applying the ‘fast-fail’ approach to evaluating κ-opioid antagonism as a treatment for anhedonia. Nat. Med. 26, 760–768 (2020).

39. Ohmura, Y. & Nagayasu, K. Functional diversity of serotonin neurons in the dorsal and median raphe nuclei in emotional responses. Neuropsychopharmacol. Rep. 45, e70015 (2025).

40. Nakamura, K. The role of the dorsal raphé nucleus in reward-seeking behavior. Front. Integr. Neurosci. 7, 60 (2013).

41. Li, Y. et al. Serotonin neurons in the dorsal raphe nucleus encode reward signals. Nat. Commun. 7, 10503 (2016).

42. Miyazaki, K., Miyazaki, K. W. & Doya, K. Activation of dorsal raphe serotonin neurons underlies waiting for delayed rewards. J. Neurosci. 31, 469–479 (2011).

43. Peters, K. Z., Cheer, J. F. & Tonini, R. Modulating the neuromodulators: Dopamine, serotonin, and the endocannabinoid system. Trends Neurosci. 44, 464–477 (2021).

44. Solomon, M. B. et al. Revisiting the role of serotonin in attention-deficit hyperactivity disorder: New insights from preclinical and clinical studies. Clin. Drug Investig. 45, 701– 742 (2025).

45. Liu, Z. et al. Dorsal raphe neurons signal reward through 5-HT and glutamate. Neuron 81, 1360–1374 (2014).

46. Qi, J. et al. A glutamatergic reward input from the dorsal raphe to ventral tegmental area dopamine neurons. Nat. Commun. 5, 5390 (2014).

47. Wang, H.-L. et al. Dorsal raphe dual serotonin-glutamate neurons drive reward by establishing excitatory synapses on VTA mesoaccumbens dopamine neurons. Cell Rep. 26, 1128–1142.e7 (2019).

48. Dayan, P. & Huys, Q. Serotonin’s many meanings elude simple theories. eLife 4, e07390 (2015).

49. Priestley, L., Mahmoodi, A., Reith, W. D., Khalighinejad, N. & Rushworth, M. F. S. Activity in human dorsal raphe nucleus signals changes in behavioural policy. Nat. Commun. 17, 1665 (2026).

50. Langley, C. et al. Computational modelling and neural correlates of reinforcement learning following three-week escitalopram: A double-blind, placebo-controlled semi-randomised study. Transl. Psychiatry 15, 175 (2025).

51. Scholl, J. et al. Beyond negative valence: 2-week administration of a serotonergic antidepressant enhances both reward and effort learning signals. PLOS Biol. 15, e2000756 (2017).

52. Stoy, M. et al. Hyporeactivity of ventral striatum towards incentive stimuli in unmedicated depressed patients normalizes after treatment with escitalopram. J. Psychopharmacol. 26, 677–688 (2012).

53. Paquelet, G. E. et al. Single-cell activity and network properties of dorsal raphe nucleus serotonin neurons during emotionally salient behaviors. Neuron 110, 2664–2679.e8 (2022).

54. Chang, A. J. et al. Neuron-type specificity of dorsal raphe projections to ventral tegmental area. Preprint at 10.1101/2021.01.06.425641 (2021).

55. Teissier, A. et al. Activity of raphé serotonergic neurons controls emotional behaviors. Cell Rep. 13, 1965–1976 (2015).

56. Gärde, M. et al. Altered serotonin 1B receptor binding after escitalopram for depression is correlated with treatment effect. Int. J. Neuropsychopharmacol. 27, pyae021 (2024).

57. Morales, M. & Margolis, E. B. Ventral tegmental area: cellular heterogeneity, connectivity and behaviour. Nat. Rev. Neurosci. 18, 73–85 (2017).

58. Root, D. H. et al. Distinct signaling by ventral tegmental area glutamate, GABA, and combinatorial glutamate-GABA neurons in motivated behavior. Cell Rep. 32, 108094 (2020).

59. Viviani, R., Abler, B., Seeringer, A. & Stingl, J. C. Effect of paroxetine and bupropion on human resting brain perfusion: An arterial spin labeling study. NeuroImage 61, 773–779 (2012).

60. Dill, T. Contraindications to magnetic resonance imaging. Heart 94, 943–948 (2008).

61. Blier, P. & De Montigny, C. Electrophysiological investigations on the effect of repeated zimelidine administration on serotonergic neurotransmission in the rat. J. Neurosci. 3, 1270–1278 (1983).

62. Henningson, C., Mlost, J. & Pollak Dorocic, I. Effects of SSRIs on the spatial transcriptome of dorsal raphe serotonin neurons. Mol. Psychiatry 10.1038/s41380-026-03644-x (2026).

63. Hamilton, M. A rating scale for depression. J. Neurol. Neurosurg. Psychiatry 23, 56–62 (1960).

64. CIPS (Collegium Internationale Psychiatriae Scalarum). Internationale Skalen für Psychiatrie [International Scales for Psychiatry] (Beltz, 1977).

65. Bagby, R. M., Ryder, A. G., Schuller, D. R. & Marshall, M. B. The Hamilton Depression Rating Scale: Has the gold standard become a lead weight? Am. J. Psychiatry 161, 2163– 2177 (2004).

66. Overall, J. E. & Gorham, D. R. The Brief Psychiatric Rating Scale. Psychol. Rep. 10, 799–812 (1962).

67. Maier, W. & Philipp, M. Comparative analysis of observer depression scales. Acta Psychiatr. Scand. 72, 239–245 (1985).

68. Franken, I. H. A., Rassin, E. & Muris, P. The assessment of anhedonia in clinical and non-clinical populations: Further validation of the Snaith–Hamilton Pleasure Scale (SHAPS). J. Affect. Disord. 99, 83–89 (2007).

69. Tzourio-Mazoyer, N. et al. Automated anatomical labeling of activations in SPM using a macroscopic anatomical parcellation of the MNI MRI single-subject brain. NeuroImage 15, 273–289 (2002).

70. Behzadi, Y., Restom, K., Liau, J. & Liu, T. T. A component based noise correction method (CompCor) for BOLD and perfusion based fMRI. NeuroImage 37, 90–101 (2007).

71. Viviani, R. A digital atlas of middle to large brain vessels and their relation to cortical and subcortical structures. Front. Neuroanat. 10, 12 (2016).

72. Viviani, R. Unbiased ROI selection in neuroimaging studies of individual differences. NeuroImage 50, 184–189 (2010).

73. World Medical Association. Declaration of Helsinki: Ethical principles for medical research involving human subjects. JAMA 310, 2191–2194 (2013).

## Supplemental References

1. Viviani, R. et al. Signals of anticipation of reward and of mean reward rates in the human brain. Sci Rep 10, 4287 (2020).

